# Sox6 expression and aneurysms of the thoracic and abdominal aorta

**DOI:** 10.1101/2022.05.23.22275278

**Authors:** David Carmona-Berrio, Isabel Adarve-Rengifo, Andrea G. Marshall, Zer Vue, Duane D. Hall, Tyne W. Miller-Fleming, Ky’Era V. Actkins, Heather K. Beasley, Paula M. Almonacid, Pierina Barturen-Larrea, Quinn S. Wells, Marcos G. Lopez, Edgar Garza-Lopez, Dao-Fu Dai, Jianqiang Shao, Kit Neikirk, Federic T. Billings, John A. Curci, Nancy J. Cox, Vivian Gama, Antentor Hinton, Jose A. Gomez

## Abstract

**Background:** Abdominal and thoracic aortic aneurysms (AAA; TAA) remain a large cause of deaths worldwide. This is in part a result of the lack of prognostic markers or early warning signs, leading to undiagnosed aortic aneurysms. Sox6 has been found to function as a regulator of renin expression controlling the rate limiting step in the renin angiotensin aldosterone system. We hypothesized that the transcription factor Sox6 may serve as an important regulator of mechanisms contributing to hypertension induced aortic aneurysms.

**Methods:** Our approach includes mRNA analysis, immunohistology staining, and protein expression studies in human samples from patients affected with AAA and TAA. In vivo, we use Angiotensin (II) to induce AAA in mice with a tamoxifen inducible Cre to specifically knock out Sox6 in smooth muscle cells. Additionally, we utilize large-scale biobank data linking de-identified medical records with genotype information to perform phenotype and laboratory-wide association scans to assess the effects of *SOX6* expression in a clinical cohort.

**Results:** In a large biobank population, *SOX6* gene expression is associated with aortic aneurysm in humans of European ancestry. Protein expression of Sox6 and TNFα was upregulated in tissue from patients affected by AAA and TAA. Moreover, we found that knocking out Sox6 in smooth muscle cells protected mice from hypertension-induced AAA, suggesting that Sox6 may be a molecular target in aortic aneurysms.

**Conclusions:** The data presented here suggest that the transcription factor Sox6 functions in the development of abdominal aortic aneurysms, and hypertension-induced rupture.

**Clinical Perspective:**

- Using electronic health records and biobank samples, we found that the transcription factor *SOX6* is associated with abdominal and thoracic aortic aneurysm and its expression is upregulated in tissue from patients affected by those diseases.
- Laboratory-wide association study (LabWAS) provides several clinical laboratory measurements associated with aortic aneurysm diagnosis that may be potential biomarkers for the disease.
- Mice with smooth muscle-specific Sox6 knock out attenuated hypertension-induced abdominal aortic aneurysm. These novel mice may be useful tools to elucidate the mechanisms associated with abdominal aortic aneurysm.

## Introduction

Abdominal aortic aneurysm (AAA) is a devastating disease that can lead to rupture of the aorta and is estimated to cause up to 200,000 yearly deaths worldwide ^1, 2^. An estimated 2.78 per 100,000 people suffer from an aortic aneurysm or dissection worldwide ^2, 3^. In the United States, aortic aneurysms are more common in men and present in 4-8% of those undergoing screening ^4, 5^. Importantly, there are no prognostic markers for AAA development, and warnings for impending aortic rupture are lacking. Amongst aortic aneurysms, abdominal aortic aneurysms are the most common, and thoracic aneurysms are less common but more heritable. For example, several connective tissue diseases are associated with the development of thoracic aortic aneurysms including Marfan syndrome (MFS), Loeys-Dietz syndrome (LDS), and Ehlers-Danlos syndrome (EDS) ^6^. Marfan syndrome is characterized for the presence of TAA caused by a mutation in fibrillin-1 ^6^. Preclinical models of TAA have identified TGFβ signaling is key in aneurysm development, and in these models, Ang II is responsible for increased TGFβ signaling ^7^. Aortic aneurysms develop when the vessel wall dilates at specific points. This process increases the risk of aorta dissection, during which the vessel intima is damaged. In the event of aorta dissection, blood enters the underlying layers of the vessel wall and create a false lumen where coagulation, immune cell infiltration, and further inflammation occur. In the case of rupture, aortic aneurysms and dissections can be very fatal.

Hypertension is a major risk factor for AAA and risk of rupture is increased by 30% for each 10-mmHg increase in blood pressure in humans ^8^. While there is compelling evidence to support the role of hypertension in AAA development ^3, 9, 10^, the pathophysiology linking them remains poorly understood. Hypertension is associated with overactivation of the renin angiotensin aldosterone system (RAAS), which facilitates oxidative stress and degradation of the extracellular matrix. This leads to smooth muscle cell death within the aorta, which may contribute to the development of aortic aneurysms^11^. In RAAS, the main effector molecule is the vasoconstrictor Angiopoietin II (Ang II), which raises blood pressure primarily through activation of type 1 angiotensin receptors (AT1R) ^12^. Ang II regulates endothelial permeability and serves as a growth factor for new blood vessels. Ang II is furthermore linked to hypertension, age-induced vascular stiffening, atherosclerosis, AAA and dissection development ^13, 14^. Here we seek to further identify the relationships linking hypertension to the development of aortic aneurysms so that additional diagnostic markers and therapeutic targets may be developed to decrease mortality and morbidity from AAA and TAA. We have previously found that Sox6 has a function as a regulator of renin expression controlling the rate limiting step in RAAS ^15, 16^. Knocking out Sox6 in Ren1d+ cells inhibited renal artery stenosis induced hypertension and kidney injury ^15^. *Sox6* has previously been reported as a specific regulon of smooth muscle cells in a single cell RNA sequence (RNAseq) study of murine abdominal aortic aneurysm ^17^. Sox6 is a transcription factor member of the D-subfamily of the sex-determining region. It is in the y-related family of transcription factors, characterized by a conserved DNA-binding domain known as the high mobility group box and an ability to bind the minor groove of DNA ^18^. Sox6 is associated with numerous disease states including cancer, cardiomyopathy, and diabetic nephropathy^19^. We hypothesized that the transcription factor Sox6 may serve as an important regulator of mechanisms contributing to aortic aneurysms, and here we present human and mouse data indicating that Sox6 has a function in the development of abdominal and thoracic aortic aneurysms.

Using a large, clinical biobank population, we found that imputed gene expression of *SOX6* is associated with aortic aneurysm in humans of European ancestry. Moreover, analysis of the expression of SOX6 in tissue from patients affected by abdominal and thoracic aortic aneurysm was upregulated and knocking out Sox6 in smooth muscle cells inhibited the development of abdominal aortic aneurysm in mice. The data presented here suggest that the transcription factor Sox6 has a previously unreported function in the development of abdominal aortic aneurysms, and that *SOX6* may be a molecular marker of human AAA and thoracic aortic aneurism (TAA).

## Methods

### Animals

Mice were housed and cared for at the Vanderbilt University Medical Center (VUMC) Division of Animal Care following the National Institutes of Health (NIH) guidelines and the Guide for the Care and Use of Laboratory Animals, US Department of Health and Human Services. All animal procedures were approved by the VUMC Institutional Animal Care and Use Committee prior to starting the experiments. All the animals used in the in vivo studies, were maintained on a 12-hour light / 12-hour dark cycle at an ambient temperature of 24°C and 60% humidity.

### Osmotic mini pump implantation

Mice were anesthetized with isoflurane for the duration of the procedure. Adult male Myh11Cre^ERT2^/Sox6^fl/fl^ (Sox6 KO) and Myh11Cre^ERT2^/Sox6^wt/wt^ (Sox6 WT) were intraperitoneally injected with 2.5 mg of Tamoxifen for five consecutive days to induced *Sox6* knock-out. One week later, mice were anesthetized using isoflurane, and osmotic mini-pumps (alzet model 2004) implanted subcutaneously for continuous infusion of Ang II (1000 ng/kg/min x 28 days) or its vehicle. A subcutaneous (SC) injection of ketoprofen (Dose: 5mg/kg BW) was given before procedure and 24 h as post-operative analgesia.

### Blood pressure measurement

Blood pressure measurements were performed using a tail-cuff method following previously reported recommendation for precise measurements previously reported ^15^. Blood pressure was also measured 4 weeks after Ang II infusion. **Human studies.** Aortic Aneurysm Aorta (AAA), Thoracic Aorta Aneurysm (TAA) and Control Aorta (Control) samples were collected from the OR at Vanderbilt University Medical Center, flash freeze in liquid nitrogen, and stored at −80°C. Full informed consent was obtained for all tissue samples. IRB #191351 “Perioperative vascular reactivity”. Approved by the Human Research Protections Program – HRPP, Vanderbilt University Medical Center. Manus J Donahue Ph.D., Chair. Institutional Review Board Health Sciences Committee #2.

### Calculating genetically-regulated gene expression (GREX) in BioVU

Vanderbilt University Medical Center curates a biorepository of genotype data matched to de-identified electronic health records (EHR) for over 259,000 individuals. Genotype data for 94,474 BioVU individuals was generated using the Illumina MultiEthnic Genotype Array (MEGAEX) and imputed to the HRC reference panel using the Michigan imputation server ^20, 21^. Genetically regulated gene expression was calculated in BioVU individuals from models built using the genotype-tissue expression (GTEx) project data ^22^ (for more details see supplemental methods).

### GREX Phenotype association study in BioVU

Aortic aneurysm cases and controls were identified in BioVU by at least two mentions of the aortic aneurysm phecode (442.1) mapped from ICD9/10 diagnosis billing codes (International Classification of Diseases, 9th and 10th editions). Controls included individuals without any mention of the aortic aneurysm phecode (see Supplemental Methods). We identified 1,208 individuals of European ancestry and 98 individuals of African ancestry as cases for aortic aneurysm in BioVU (controls = 58,099 and 13,551, respectively). Logistic regression was used to examine the relationship between *SOX6* GREX or AAA/TAA gene GREX and the aortic aneurysm disease outcome. Age, sex, median age of medical record, principal components 1-10, genotype batch, and number of medical center visits were used as covariates in the analysis. More details in Supplemental Methods.

### GREX Laboratory-wide association scan (LabWAS) in BioVU

We evaluated all labs with measurements from at least 100 individuals, which resulted in testing 323 labs across 70,337 individuals of European ancestry and 241 labs across 15,123 individuals of African ancestry. We examined the relationship between the GREX of *SOX6* and additional AAA/TAA genes with all available clinical lab measurements in linear regression models. Age, sex, principal components 1-10, genotype batch, median age of medical record, and number of medical center visits were used as covariates in the analysis. We used a Bonferroni-corrected threshold to account for the number of GREX-lab pairs present in the associations tested (i.e. 0.05/(number of labs*number of unique gene-tissue pairs)). More details in Supplemental Methods.

### Aortic aneurysm diagnosis LabWAS in the Vanderbilt Synthetic Derivative

The Vanderbilt Synthetic Derivative includes insurance billing codes (International Classification of Diseases, 9^th^ and 10^th^ editions/ICD-9 and ICD-10 codes), laboratory measurements, and clinician notes. We used the Quality Labs pipeline to extract lab measurements in individuals classified as cases or controls for aortic aneurysm based on the phecode 442.1 for aortic aneurysm. We evaluated 335 labs across 2,273 cases and 1,268,204 controls for aortic aneurysm. Age, sex, median age of medical record, EHR-reported race and ethnicity, and number of medical center visits were used as covariates in the analysis. We used a Bonferroni-corrected threshold to account for the number of labs tested (i.e. 0.05/(number of labs tested)). For more details see Supplemental Methods.

### Statistics

Two-way ANOVA was used for experiments with three or more conditions followed by Tukey’s tests for comparisons between individual groups. One way ANOVA was used for experiments with three conditions. Student t-test was used to compare the mRNA expression values between control aortic sample and TAA or control aortic sample and AAA. A p-value equal or less than 0.05 was considered significant. All statistical analysis were performed using GraphPad Prism 8.2.

## Results

To determine the relationship between *SOX6* and aortic aneurysms in humans, we modeled genetically regulated gene expression (GREX) of *SOX6* across 70,439 individuals of European ancestry and 15,174 individuals of African ancestry in the Vanderbilt biobank, BioVU (Figure 1). Individuals diagnosed with aortic aneurysm were identified using phecodes mapped from ICD9/10 billing codes from de-identified electronic health records. The relationship between *SOX6* GREX and the aortic aneurysm diagnosis was then evaluated by logistic regression models, correcting for the age, sex, and genetic ancestry of each individual. In a large sample of BioVU individuals of European ancestry, we found that *SOX6* GREX is significantly associated with aortic aneurysm diagnosis (n=1,208, p=0.0265, Table 1). However, there was no significant association between *SOX6* GREX and aortic aneurysm in individuals of African ancestry, perhaps due to the smaller sample size (n=98, p=0.1452, Table 1).

**Figure 1.**
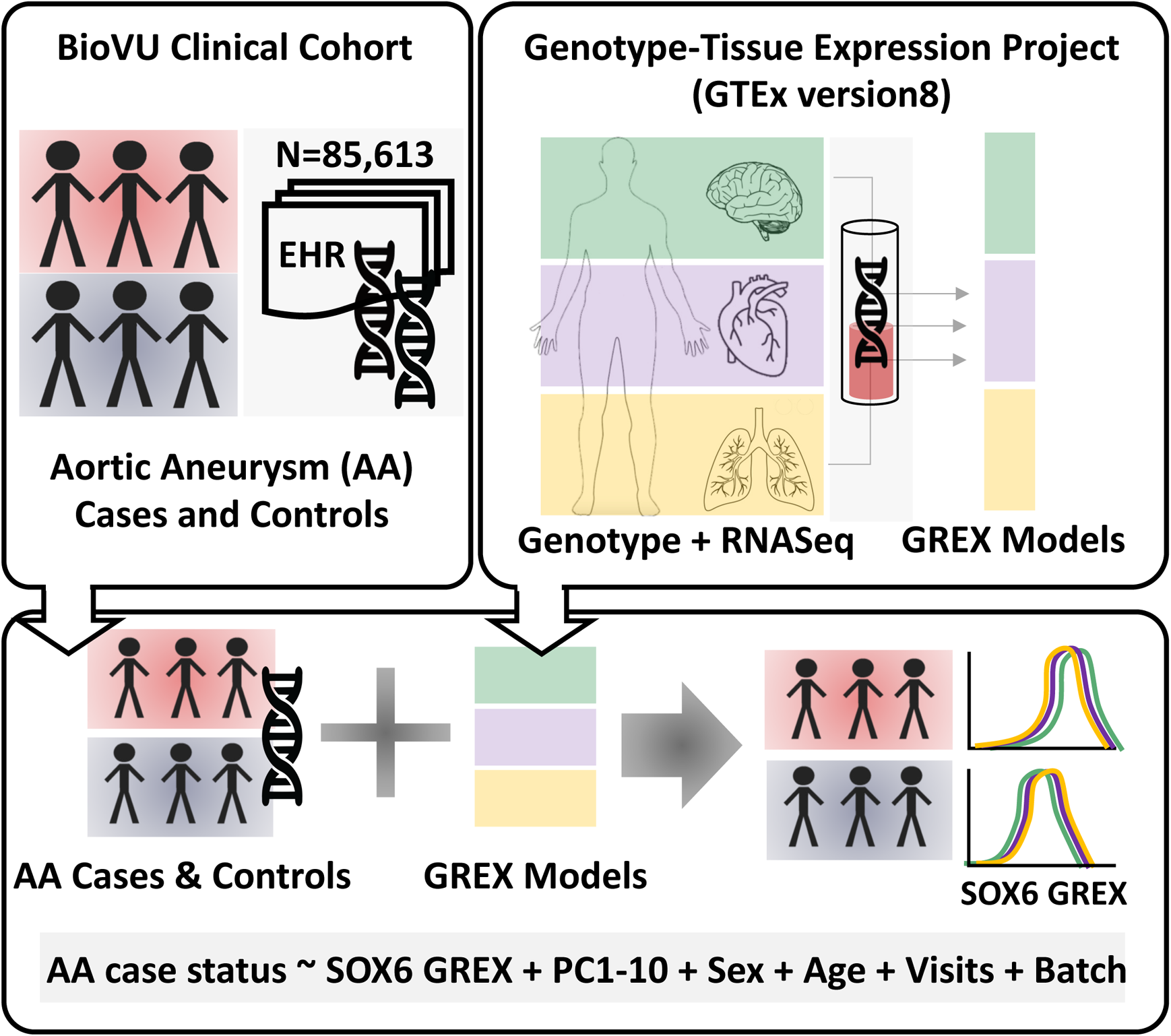
Modeling *SOX6* genetically regulated gene expression (GREX) and aortic aneurysm in a medical biobank. We identified cases and controls for aortic aneurysm (AA) from BioVU participants using ICD9/10 codes from Vanderbilt’s de-identified electronic health record database (n= 1,306 and 71,650, respectively, top left panel). Genetically regulated gene expression for *SOX6* and *SOX6*-regulated genes were calculated in BioVU participants using models built from the GTEx version 8 data (top right panel), which contains genotype data matched to RNA-Seq data from 838 donors across forty-nine tissues. Imputed gene expression was calculated and assessed for association in AA cases versus controls using logistic regression models (bottom panel), accounting for genetic ancestry (principal components/PC 1-10), sex, age, number of medical center visits, and genotyping batch.

**Table 1.** Phenotype Association Results for SOX6 GREX

| **Genetic Ancestry Group** | **Phenotype** | **GREX** | **p** | **n_models** | **n_samples** | **p_best** | **best_tissue** |
| --- | --- | --- | --- | --- | --- | --- | --- |
| **European** | **AorticAneurysm** | **SOX6** | **0.0265** | **14** | **59307** | **0.0326** | **SkinSunExposed** |
| African | AorticAneurysm | SOX6 | 0.1452 | 14 | 13649 | 0.0300 | ColonSigmoid |

Relationship between SOX6 and aortic aneurysms in humans. Genetically regulated gene expression (GREX) of SOX6 across 59,307 individuals of European ancestry and 13,649 individuals of African ancestry in the Vanderbilt biobank, BioVU.

Next, we analyzed the expression patterns of *SOX6* in human with aortic aneurysms. Tissue from patients affected by AAA and TAA were collected during aneurysm repair surgeries, and nonaneurysmal aortic tissue samples from ascending aorta punch sites for saphenous vein grafts from patients undergoing coronary artery bypass surgery were collected as a control. The burden of AAA increases with age and is more prevalent in men than women ^3^. For TAA patients, age ranged from 48 to 81 years old (Supplemental Table 1). In comparison, AAA patients ages range from 60 to 67 years old (Supplemental Table 1). Elastin staining analysis showed that in aneurysmal tissue from both AAA and TAA patients, there were significant elastic fragmentation and cystic medial degeneration (Supplemental Figure 1). Furthermore, there was loss of smooth muscle cells, increased collagen deposition, and fibrosis (Supplemental Figures 2 and 3).

To better understand the transcriptional pathways activated in disease states, we analyzed gene expression using RNAseq analysis. RNAseq was used to determine the transcriptional differences between TAA tissue and nonaneurysmal aortic tissue. RNAseq analysis shows that *SOX6* expression is upregulated in TAA (Figures 2A and B). Other genes upregulated in TAA tissue included Gremlin 1 (*GREM1*), C-C motif chemokine ligand 21 (*CCL21*), and FYN binding protein 1 (*ADIPOQ*) (Figure 2A and B). *GREM1* is a bone morphogenic protein (BMP) antagonist, upregulated in aortic tissue from Loeys-Dietz syndrome ^23^. *CCL21* was found in the conditioned medium isolated from AAA split arterial layer ^24^. Adiponectin (*ADIPOQ*) has an inhibitory function in abdominal aortic aneurysm development induced by Ang II and beta Aminopropionitrile ^25^. Ingenuity Pathway Analysis of TAA samples showed that there was downregulation of several functional pathways, including endocytosis (Figure 2C). GSEA:KEGG (Gene Set Enrichment Analysis: Kyoto Encyclopedia of Genes and Genomes) analyses indicate that in TAA, PPAR signaling and cytokine-cytokine receptor interactions are upregulated (Figure 2D). This aligns with previous studies which have found that PPARγ agonist decreases inflammation in patients with aortic aneurysm and has a function in AAA development ^26^. Ingenuity Pathway Analysis of causal networks showed that *PRKAA2*, *SGK3*, and *CNGA3* are upregulated in the TAA samples. *PRKAA2* (Protein Kinase AMP-Activated Catalytic Subunit Alpha 2) is a gene essential in energy-sensing enzyme that monitors cellular energy status, *SGK3* (Serum/Glucocorticoid Regulated Kinase Family Member 3), this gene has a role in neutral amino acid transport and activation of potassium and chloride channels, and *CNGA3* (Cyclic Nucleotide Gated Channel Subunit Alpha 3) is a gene needed for normal vision and olfactory signal transduction. We found that these genes are upregulated in human TAA samples (Figure 2E).

**Figure 2.**
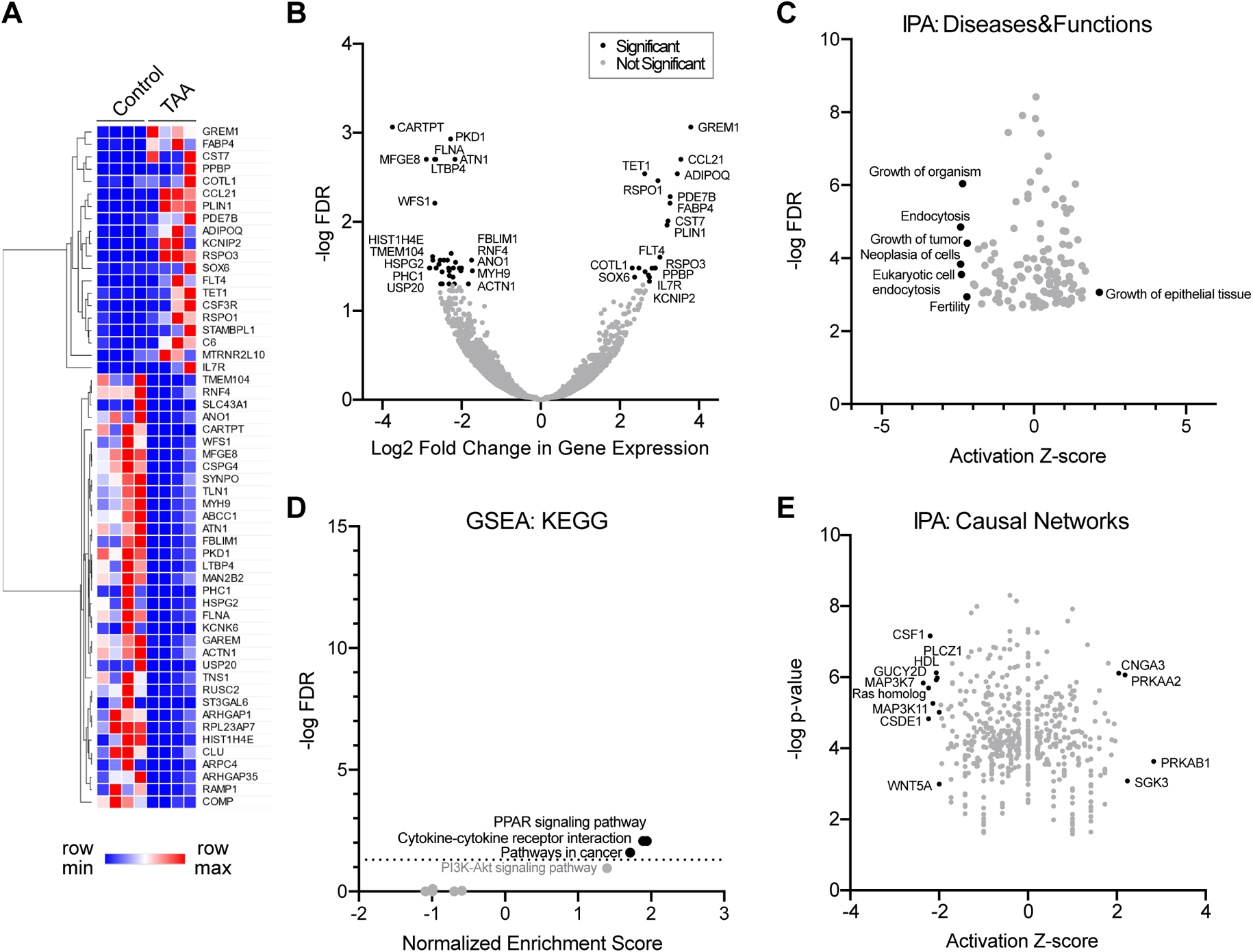
RNA sequence analysis of human TAA samples. RNA-sequencing analysis indicates a modest change in growth related pathway in thoracic aortic aneurysm (TAA) samples. **(A)** Heatmap of 55 significant differentially expressed genes (DEGs) found in TAA samples relative to control (n=4 per group). **(B)** Expression of significant (black circles) versus non-significant (gray circles) DEGs displayed as a volcano plot. FDR, false discovery rate. **(C)** Ingenuity pathway analysis (IPA) results of DEGs for enriched diseases and functions annotations. **(D)** Enriched KEGG pathway terms from gene set enrichment analysis (GSEA). **(E)** Enriched IPA causal cellular networks.

Gene transcriptional differences between AAA tissue and nonaneurysmal aortic tissue was determined using RNAseq (Figure 3A). *SOX6* gene fold change was −0.589 (p=0.53), protein expression analysis is presented later. The immune-related gene *JCHAIN* (joining chain of multimeric IgA and IgM) and the guanine nucleotide exchange factor *DENND1C* (DENN domain containing 1C) had the highest fold increase in AAA (Figure 3B). IPA showed that several canonical pathways previously associated with AAA development ^27–30^ are upregulated in AAA samples, including Th2 and Th1 pathways, PTEN signaling, and oxidative stress in macrophages (Figure 3C). Most of the upregulated functions identified by IPA are related to immune responses known to play a role in AAA ^27–30^ (Figure 3D and Supplementary Figure 4). Furthermore, IPA analysis of causal networks identified *CoREST*, *SOX11* and *ACE2* upregulated in the AAA samples (Figure 3E). *CoREST* determining neural cell differentiation, *SOX11* is important for regulation of embryonic development and in the determination of the cell fate, and consistent with our data, *ACE2* was previously reported as a modulator of angiotensin II induction of AAA ^31^.

**Figure 3.**
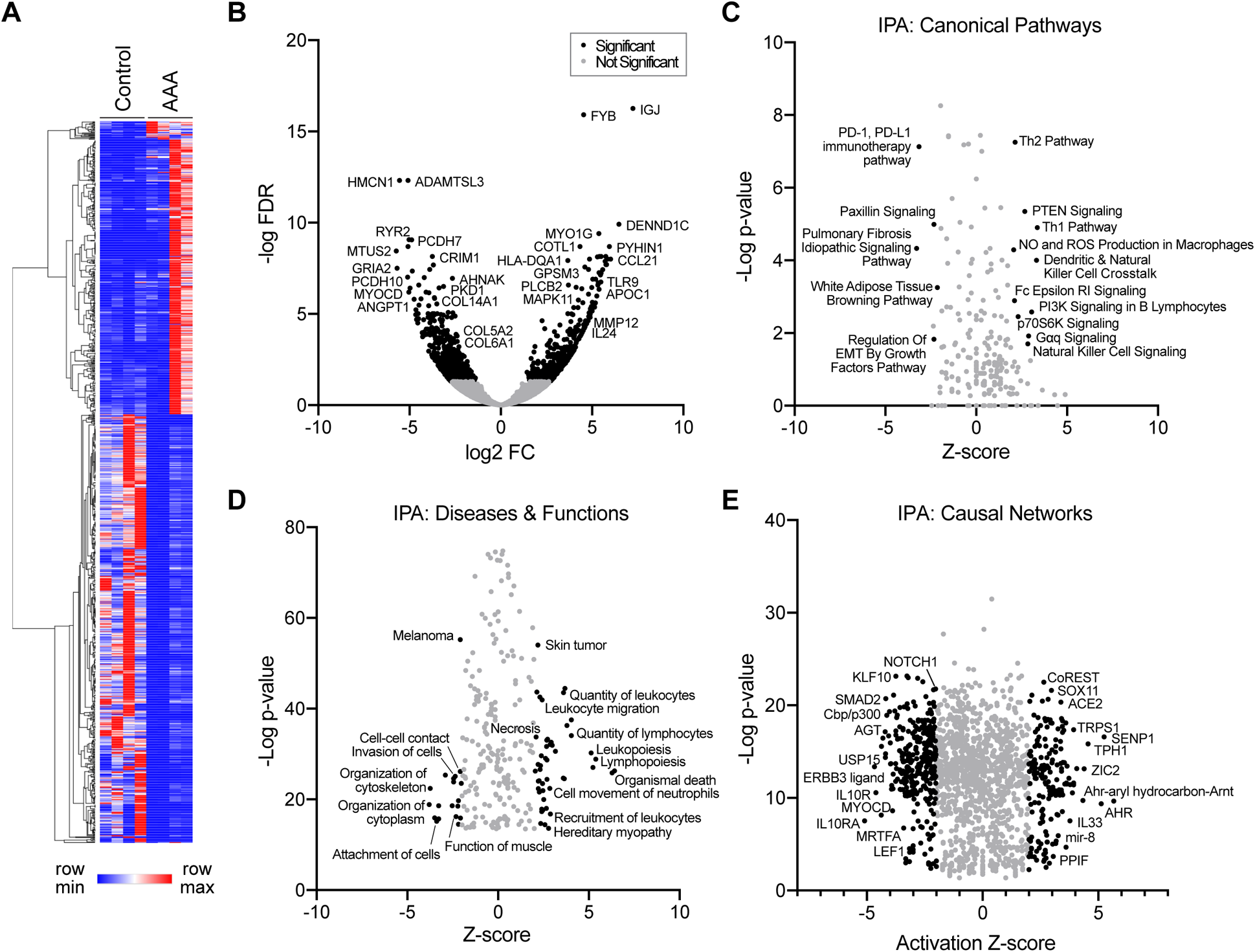
RNA sequence analysis of human AAA samples. RNA-sequencing analysis suggests a robust immune response occurs in abdominal aortic aneurysm (AAA). **(A)** Heatmap of 1022 significant differentially expressed genes (DEGs) found in AAA samples relative to control (n=4 per group). **(B)** Expression of significant (black circles) versus non-significant (gray circles) DEGs displayed as a volcano plot. Log2 FC, log2 fold change in control vs. AAA expression. FDR, false discovery rate. C-E. Ingenuity pathway analysis (IPA) results of DEGs for enriched canonical pathway **(C)**, diseases and functions annotation **(D)**, and causal network **(E)** terms. Example enriched terms are indicated.

Next, we validated the expression of genes found in the RNAseq analysis that were common to both AAA and TAA (Supplemental Figure 5 and Supplemental Table 2) whose promoters contain SOX6 binding sites using RT-qPCR (Supplemental Figure 6 and Supplemental Tables 3 and 4). RNAseq genes upregulated *COTL1*, and *CCL21* and downregulated *PKD1* in both TAA and AAA. *In silico* analysis indicates that SOX6 has binding sites in the promoter of those 3 genes, suggesting a regulatory function (Supplemental Figure 7). The *COTL1* was significantly upregulated in AAA compared to the control (Supplemental Figure 6A). *JCHAIN* expression was upregulated in AAA samples based on our RNAseq analysis. The promoter of this gene has several binding sites for SOX6, and the mRNA was upregulated in AAA samples (Supplemental Figure 6B). *SOX6* transcripts were elevated in about half of AAA and TAA samples (not significant due to small sample size (Supplemental Figure 6C). Likewise, *PKD1*, and *CCL21* were also detected in the samples (not significant, Supplemental Table 3 and 4), there were no statistically significant differences.

We then examined protein expression in human aortic samples by Western blot. SOX6 protein expression was significantly upregulated in AAA and TAA aortic tissue compared to control tissue (Figure 5A-F and Supplemental Figure 8). Tumor necrosis factor alpha (TNFα) has a known function in the development of both TAA ^32^ and AAA ^33, 34^, and is used as a positive control. The expression of TNFα was upregulated in both AAA and TAA (Figure 5A-F and Supplemental Figure 8). *In silico* analysis of the TNFα promoter shows that there are several SOX6 binding sites suggesting a possible function of SOX6 in the control of TNFα expression during both AAA and TAA development. Moreover, *Sox6* was identified as part of a regulon in murine aortic smooth muscle cells in a mouse model of AAA ^17^. These results suggest that SOX6 is upregulated in AAA and TAA, indicating a likely role in the etiology of these diseases, in line with our mouse models data.

**Figure 5.**
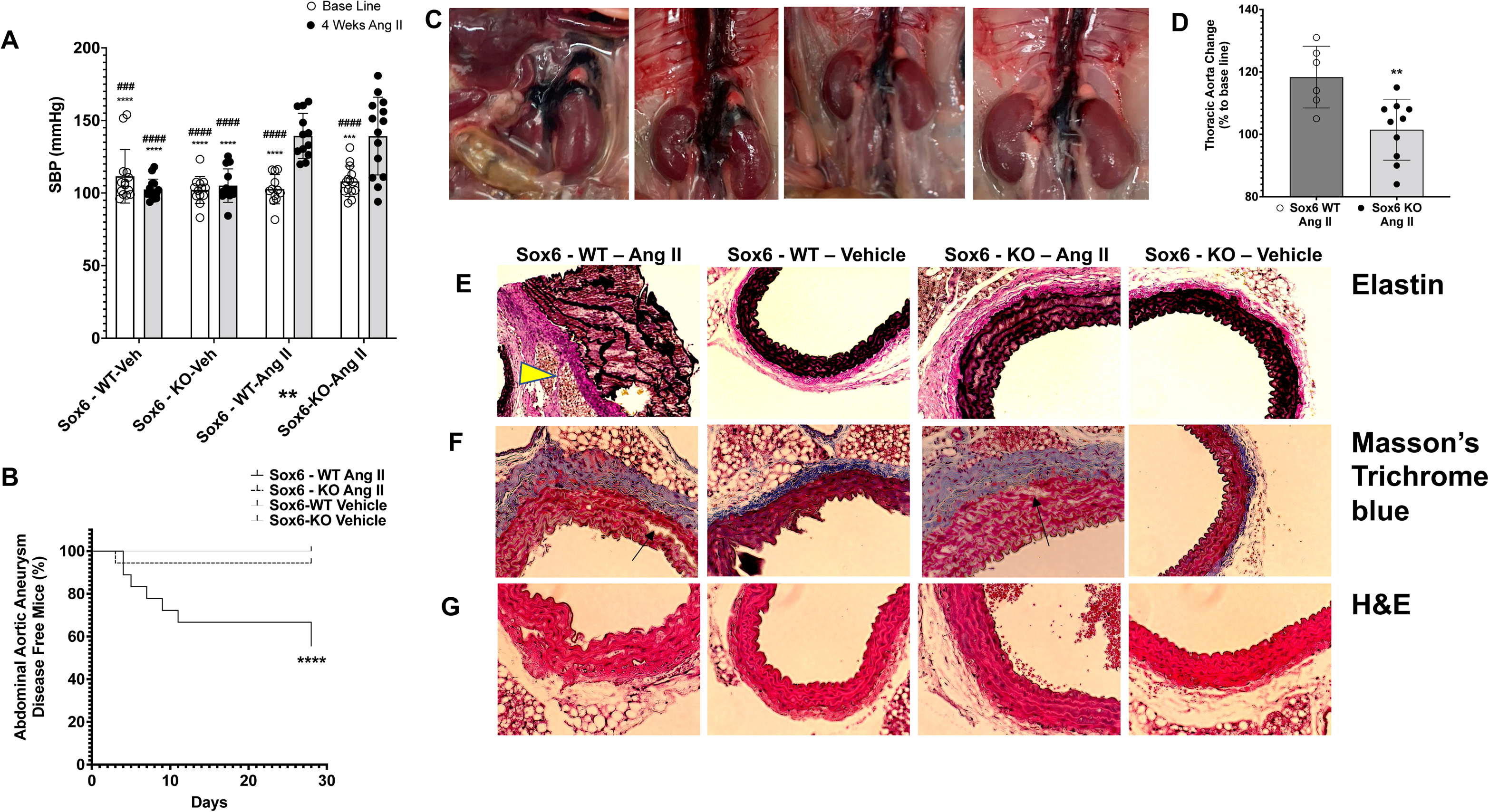
Knocking out Sox6 in smooth muscle cells inhibits hypertension induced abdominal aortic changes in mice. C57Bl6 Myh11Cre^ERT2^/Sox6^fl/fl^ (Sox6 KO) and Myh11Cre^ERT2^/Sox6^wt/wt^ (Sox6 WT) mice were intraperitoneally injected with 2.5 mg of tamoxifen for five consecutive days. Then osmotic mini pumps were implanted subcutaneously for infusion of angiotensin II (1000 ng/kg/min x 28 days). Knocking out Sox6 in SMCs inhibits degradation of ECM. **(A)** Blood pressure changes were measured using tail cuff method (n=12 to 14). P values calculated using a two-way ANNOVA. **** represents P <0.0001 when comparing Sox6 KO Ang II treated group to both vehicles. #### represents P < 0.0001 when comparing Sox6 WT Ang II treated group to both vehicles. **(B)** Ang II induced abdominal aortic aneurysm in Sox WT mice (n=18). P values calculated using log-rank (Mantel-Cox) test. Sox6 WT vehicle (n=14), Sox6 KO vehicle (n=13), and Sox6 KO Ang II (n=18). *** represents P < 0.001. **(C)** Images of 4 representative Sox6 WT mice died of AAA rupture. **(D)** Ultrasound measurement of thoracic aortic diameter in mice presented as the ratio of post / before Ang II. Sox6 WT N=6. Sox6 KO N=10. P values calculated using unpaired T - test. ** represents P < 0.01. Representative images. 20X magnification. **(E)** Van Gieson Stain for elastin showed cystic medial degeneration leading to aneurysm formation in Sox6 WT - Ang II (yellow arrowhead). **(F)** Trichrome blue showed areas of elastic fragmentation in both Sox6 WT - Ang II and Sox6 KO - Ang II (arrows). **(G)** Hematoxylin and Eosin.

We sought to evaluate the clinical implications of genes upregulated in the TAA and AAA tissues by leveraging the large medical biobank, BioVU. The GREX for the genes identified by TAA and AAA tissue RNAseq analysis was calculated in BioVU participants (Table 2 and 3 show Patient Demographics). Additionally, clinical laboratory measurements were extracted for 70,337 individuals of European ancestry and 15,123 individuals of African ancestry (323 and 241 lab tests, respectively) using the Quality Lab pipeline (Supplemental Figure 9)^5^. We tested the relationship between AAA and TAA gene GREX (*ADIPOQ*, *APOC1*, *COTL1*, *CST7*, *DENND1C*, *FABP4*, *FLT4*, *GREM1*, *HLADQA1*, *IL7R*, *MYO1G*, *PKD1*, *PLIN1*, *PYHIN*1, *RSPO3*, *TGFB1*, *TLR9*) and lab measurements using linear regression models, correcting for age, sex, and genetic ancestry across all patients. We found significant associations between *APOC1* (Apolipoprotein C1) GREX with triglyceride in serum or plasma (n=32,852, p=2.11×10^−14^) and *HLA-DQA1* (Major Histocompatibility Complex, Class II, DQ Alpha 1) GREX with glycated hemoglobin A1c (n=23,162, p=1.31×10^−13^), glucose (n=66,124, p=8.19×10^−8^), and tau protein in body fluid (n=47,235, p=3.29×10^−6^) across individuals of European ancestry (Supplemental Figure 10, Supplemental Table 5). *APOC1* was reported as a possible molecular marker for AAA ^35^. A locus in the HLA-DQA1 gene was identified as a possible risk factor of AAA development ^36^. We did not note any significant associations between GREX and lab values in the much smaller sample of individuals of African ancestry (Supplemental Figure 11, Supplemental Table 6).

**Table 2.** Demographics of Patients with Atrioventricular Block

| Demographics | Case | Control | p |
| --- | --- | --- | --- |
| N | 3,812 | 427,662 |  |
| Mean Age (SD) | 68.92 (15.11) | 46.02 (16.96) | <0.001 |
| Mean BMI (SD) | 29.01 (6.18) | 28.84 (6.82) | 0.114 |
| Race (%) |  |  | <0.001 |
| White | 3,439 (90.2) | 346,245 (81.0) |  |
| Black | 285 (7.5) | 47,912 (11.2) |  |
| Asian | 29 (0.8) | 8,254 (1.9) |  |
| Other | 59 (1.5) | 25,251 (5.9) |  |
| Sex (%) |  |  | <0.001 |
| Female | 1,609 (42.2) | 270,107 (63.2) |  |
| Male | 2,203 (57.8) | 157,549 (36.8) |  |
| Unknown | 0 (0.0) | 6 (0.0) |  |
| Ethnicity (%) |  |  | <0.001 |
| Hispanic | 20 (0.5) | 11,240 (2.6) |  |
| Non-Hispanic | 3,752 (98.4) | 395,374 (92.5) |  |
| Unknown | 40 (1.0) | 21,048 (4.9) |  |

Other includes individuals with an EHR-reported race of Asian, Native American, Pacific Islander, Indian, other, mixed race, unknown, and did not report. BMI = body mass index

**Table 3.** Demographics of Patients with Chronic Vascular Insufficiency of Intestine

| Demographics | Case | Control | p |
| --- | --- | --- | --- |
| N | 3,812 | 427,662 |  |
| Mean Age (SD) | 59.88 (17.42) | 47.17 (17.40) | <0.001 |
| Mean BMI (SD) | 25.44 (5.55) | 28.97 (6.90) | <0.001 |
| Race (%) |  |  | 0.04 |
| White | 283 (86.8) | 435,313 (81.4) |  |
| Black | 29 (8.9) | 60,824 (11.4) |  |
| Asian | 6 (1.8) | 9,648 (1.8) |  |
| Other | 8 (2.5) | 29,144 (5.4) |  |
| Sex (%) |  |  | 0.14 |
| Female | 221 (67.8) | 333,960 (62.4) |  |
| Male | 105 (32.2) | 200,962 (37.6) |  |
| Unknown | 0 (0.0) | 7 (0.0) |  |
| Ethnicity (%) |  |  | 0.02 |
| Hispanic | 5 (1.5) | 13,330 (2.5) |  |
| Non-Hispanic | 316 (96.9) | 497,857 (93.1) |  |
| Unknown | 5 (1.5) | 23,742 (4.4) |  |

Other includes individuals with an EHR-reported race of Asian, Native American, Pacific Islander, Indian, other, mixed race, unknown, and did not report. BMI = body mass index

In follow-up analyses, we tested the association between the AAA and TAA genes identified by RNASeq analysis with aortic aneurysm diagnosis in the BioVU patient population (Figures 2 and 3). We modeled gene expression in the multiple genes identified by RNASeq analysis (*ADIPOQ*, APOC1, *COTL1*, *CST7*, *DENND1C*, *FABP4*, *FLT4*, *GREM1*, *HLADQA1*, *IL7R*, *MYO1G*, *PKD1*, *PLIN1*, *PYHIN1*, *RSPO3*, *TGFB1*, *TLR9*) and found a nominal association with ADIPOQ GREX in individuals of European ancestry (p = 0.0455) with aortic aneurysm diagnosis (Supplemental Table 7). Adiponectin (*ADIPOQ*) has an inhibitory function in abdominal aortic aneurysm development induced by Ang II and beta Aminopropionitrile ^25^.

To determine if Sox6 plays a key role in the development of hypertension-induced AAA, we generated a novel mouse model: smooth muscle cell specific Sox6 specific knock out using a tamoxifen inducible Cre, the Myh11^CreERT2^/Sox6^fl/fl^ (Sox6 KO) mice. Myh11^CreERT2^/Sox6^wt/wt^ (Sox6 WT) littermate mice were used as controls ^37^. Cre expression was induced with tamoxifen intraperitoneal injections for five consecutive days to modify Sox6 expression prior to Ang II. Sox6 WT and Sox6 KO adult mice received continuous infusion of Ang II 1000 ng/kg/min for 28 days in a subcutaneous Alzet 2004 pump. Tamoxifen did not affect blood pressure or induce AAA. The baseline SBP were similar across all groups (102.7 ± 10.1 mmHg in Sox6 WT and 108.1 ± 10.5 mmHg in Sox6 KO): Ang II increased systolic blood pressure by about 35 mmHg (139.4 ± 15.5 in Sox6 WT and 139.2 ± 26.8 in Sox6 KO) (Figure 5A). Kaplan Meier curves demonstrate that 44 % of the Sox6 WT mice died of AAA rupture, most within the initial 11 days of Ang II infusion, in contrast to only 6% mortality in Sox6 KO mouse (Log-rank (Mantel-Cox) test p<0.001, Figure 5B and C). AAA did not develop in control vehicle-treated mice (Figure 5B). Figure 5C shows representative images of Ang II-induced fatal rupture of AAA in Sox6 WT mice. Using ultrasound measurement, we showed that Ang II induced dilation of thoracic aortas is significantly higher in Sox6 WT than those in Sox6 KO (Figure 5D). Histological analysis of the abdominal aortas of the surviving mice shows cystic medial degeneration leading to AAA in Sox6 WT - Ang II (Figure 5E). Elastin fragmentation and degradation were observed in Ang II treated mice for both genotypes (Figure 5E), but were absent in the vehicle-treated controls, suggesting that Ang II induced elastic fragmentation, cystic medial degeneration, leading to aortic dilation and AAA development (Figure 5C-G) Taken together, our data suggest that Sox6 controls a smooth muscle cell transcription network important in the development of AAA induced by hypertension and knockout of Sox6 in smooth muscle cells may de-couple hypertension from AAA development.

## Discussion

Here we utilize multiple approaches, including a model of abdominal aortic aneurysm in Sox6 knockout mice by Ang II induced hypertension, human clinical health records linked to genotype data, and mRNA and protein expression in human samples from patients affected with abdominal and thoracic aortic aneurysm to evaluate the relationship between *SOX6* and aortic aneurysm. The results of this report establish that the transcription factor Sox6 has a new function in the development of hypertension induced abdominal aortic aneurysm. Moreover, analysis of human clinical data indicates that *SOX6* is associated with both abdominal and thoracic aortic aneurysms.

There is compelling evidence to support the role of hypertension in AAA development ^3, 8–10^. However, current antihypertensive therapies do not typically work against aortic aneurysm growth and rupture ^38^, with success only observed in patients suffering aortic aneurysm due to Marfan syndrome ^39^. Our data indicates that knocking out Sox6 in smooth muscle cells (Sox6 KO) decouples hypertension and AAA development (Figure 5). Sox6 WT mice that develop AAA rupture died at about one-third of the treatment time and account for 44% of the population treated with Ang II. This indicates that in our animal regiment that includes tamoxifen induction of Cre expression, more than one-third of the Myh11^CreERt2^/Sox6^wt/wt^ - wild type for Sox6 are susceptible to hypertension-induced AAA rupture. The ECM organization of the abdominal aorta in the Sox6 KO mice was preserved from hypertension-induced damage (Figure 5E-G), indicating that smooth muscle cells are key in the Ang II induced ECM damage with Sox6 playing a key function. Additionally, we find that *SOX6* GREX is significantly associated with hypertension diagnosis in BioVU and within the UK Biobank dataset (Supplemental Table 8). Gene centric arrays and genome wide association studies (GWAS) indicate a strong association of Sox6 with hypertension (reviewed in ^19^). In combination, these data suggest that Sox6 has a novel function in the development of aortic aneurysms and may serve as a promising target.

Electronic health records linked to patient genotype data aid in the discovery of new markers of human diseases and soon will aid in the development of personalized medicine approaches and therapies. Using the institutional resource BioVU we found that imputed expression of *SOX6* is associated with aortic aneurysm diagnosis. Moreover, expression of this transcription factor was upregulated in both abdominal and thoracic aortic aneurysm tissue samples, and in RNAseq from TAA samples *SOX6* was one of the genes upregulated, indicating a direct transcriptional effect of *SOX6* in these diseases. Furthermore, in TAA samples, upregulation of genes involved PPAR signaling, and cytokine receptor interaction are upregulated in TAA samples indicating a key role for the immune system in the disease of this stage (Figure 2C and D). Previous reports indicate that PPARγ has a function in AAA development ^26^.

Analysis of aneurysmal aortic tissue from patients with AAA by RNAseq indicates that in AAA there are several canonical pathways upregulated that involved the immune system, oxidative damage, macrophages production of NO and reactive oxygen species (ROS), natural killer cells signaling, and Th2 pathway (Figure 3C). The disease pathways upregulated were immune system driven such as leukocyte migration and quantity, leukopoiesis, and lymphopoiesis (Figure 3D). There are differences in the AAA and TAA RNAseq that may reflect molecular distinctions in the development of both diseases. These results confirm the key role the immune system plays in the development of AAA ^27–30^. The AAA tissue samples were compared to nonaneurysmal aortic samples collected from patents undergoing coronary artery bypass surgery and as such our findings may reflect a partial assessment of the genes upregulated in the AAA samples. However, in the mouse model we found that the elastin in the abdominal aorta of Sox6 WT mice is disarranged, and that there is an increase in fibrosis, as seen in the human samples (Figure 5E-G and Supplemental Figures 1 to 3).

AAA accounts for more than 175,000 deaths globally with AAA rupture causing 1% of deaths in men aged over 65 years ^8^. Transabdominal ultrasound is used to diagnose AAA, patients 65 years or older with a history of smoking should be screened with at least one ultrasound at 65 years old and the detection by rupture is not common. The disease is usually detected upon rupture, carrying an 80% mortality rate ^4, 40^. TAA symptoms are rare, making it difficult to diagnose, ^41^ with a high percentage of TAA patients dying without medical attention ^41, 42^. Laboratory tests performed at hospitals and clinics are widespread practices to diagnose diseases and these lab results are reported in the electronic health record (EHR). The BioVU database contains de-identified EHRs linked to patient genotype data. One of the challenges of the treatment for thoracic and abdominal aortic aneurysm is the asymptomatic nature of both diseases. TAA is harder to diagnose ^41^, requiring cross sectional imaging (CT or MR), and there is a lack of molecular markers for predicting disease progression; therefore, it is important to develop molecular markers and clinical tests to better diagnose TAA and AAA. BioVU links genomic and clinical data from a large sample of patients and provides the opportunity to identify clinical data that is associated with these diseases. To address this, we performed a laboratory-wide association study (LabWAS), using the BioVU resource to identify the laboratory measurements associated with an aortic aneurysm diagnosis (Supplemental Figure 12, Supplemental Table 9). Aortic aneurysm diagnosis was significantly associated with multiple biomarkers and measurements of cardiovascular and metabolic health, including cholesterol, glucose, calcium, hemoglobin A1C, triglycerides, QRS duration, T-wave axis from electrocardiograms. Additionally, we see significant associations to creatinine, immune system markers (white blood cells, eosinophils, lymphocytes), and red blood cell width and distribution.

LabWAS results (Supplemental Figure 12, Supplemental Table 9) indicate that changes in erythrocytes distribution width is associated with aortic aneurysms. Erythrocytes wide distribution association with aortic aneurysms is reported ^43^ and based in our findings may be a good clinical laboratory test to detect aortic aneurysm. The clinical cardiac parameters QRS duration and ST segment ware associated with aortic aneurysm in our analysis. Two reports suggest that QRS may serve as an additional risk assessment of aortic dissection ^44^, and ST segment depression concomitant with T-wave inversion was used to diagnosed type A aortic dissection in a hypertensive patient ^45^. Decrease in creatinine clearance associated with increase serum creatinine after endovascular aneurysm repair was reported after a meta-analysis ^46^. Our LabWAS found creatinine as a parameter associated with aortic aneurysms. High-density lipoprotein (HDL) and calcium clinical test are strongly associated with aortic aneurysms. Low HDL is considered a risk factor for AAA ^47^. Other clinical parameters found in Supplemental Figure 12, Supplemental Table 9 may serve a clinical test to detect aortic aneurysm aiding in the diagnosis of AAA and TAA.

Additionally, we performed a LabWAS within the BioVU population to discern associations between genetic variation across *SOX6* and *SOX6*-regulated genes. There are significant associations between APOC1 GREX with triglyceride in serum or plasma, and HLA-DQA1 GREX with glycated hemoglobin A1c, glucose, and tau protein in individuals of European ancestry (Supplemental Figure 9, Supplemental Table 5). *APOC1* and *HLA-DQA1* were reported as possible molecular markers of AAA ^35, 36^. Our data and previous reports ^23, 25, 48^ indicate that Adiponectin may serve as a molecular marker for aortic aneurysm in patients of European ancestry. No significant results were detected in patients of African ancestry likely due to the smaller sample size (Supplemental Figure 11, Supplemental Table 6). Analysis within BioVU suggests that *SOX6* may represent a new molecular marker for AAA and TAA. The use of clinical laboratory measurements in the EHR may benefit translational research; however, the utility of this type of analysis is yet to be determined ^49^ and no conclusive results are reported yet. However, our findings provide multiple clinical lab values associated with aortic aneurysm diagnosis, providing a more comprehensive look at potential biomarkers for the disease. These results may promote the development of improved patient risk stratification for aortic aneurysms and may lead to more informed referral for imaging for definitive diagnosis of aortic aneurysms.

In consensus with other reports, TNFα protein expression is upregulated in human TAA tissue ^32, 50^, and its expression is also increased in human AAA tissue (Figure 4). Concomitant with TNFα, the expression of SOX6 with significantly upregulated in both human thoracic and abdominal aortic aneurysm tissue suggesting a function for SOX6 in the transcription of genes associated with these diseases. The human tissue use in our study was collected from patients affected by AAA and TAA of thoracic and abdominal aorta once the aneurysm is developed, indicating that *SOX6* plays a role in the advanced disease stage in humans.

**Figure 4.**
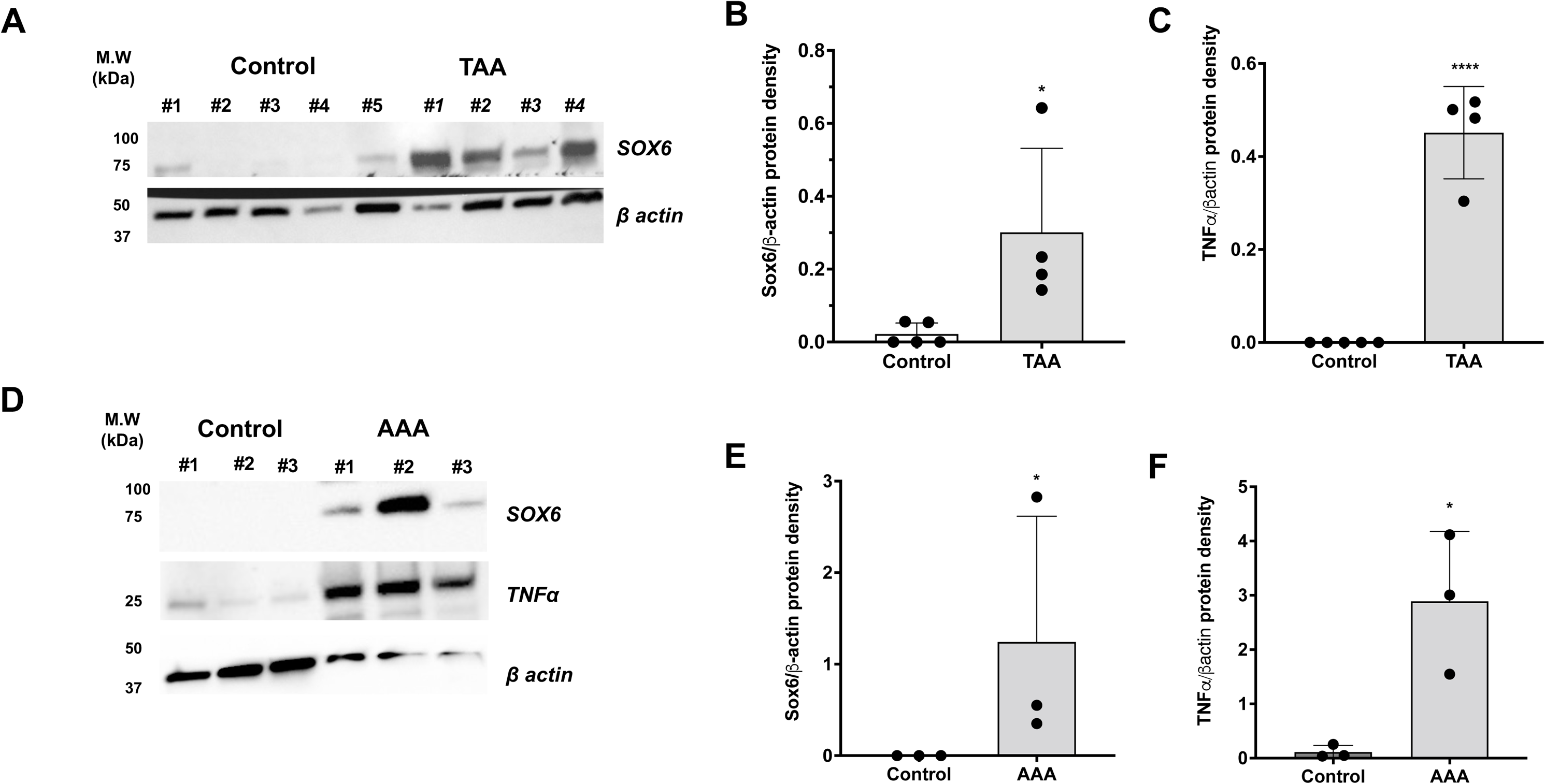
Human abdominal and Thoracic aortic aneurysm Sox6 expression Western blot analysis. SOX6 expression increases in human AAA and TAA samples. Human aorta samples were collected from tissue discarded after AAA or TAA or coronary artery bypass surgery (control). Samples were flash freeze in liquid nitrogen and stored at −80°C. Expression of SOX6 was analyzed by Western blot using a specific antibody. **(A)** representative Western blots images showing levels of SOX6 expression in human TAA. **(B)** Densitometric analysis of SOX6 and TNFα protein bands show right of Western blots. **(C)** representative Western blots images showing levels of SOX6 and TNFα expression in human AAA. Beta-actin was used as a loading control. **(D)** Densitometric analysis of SOX6 and TNFα protein bands show right of Western blots. TAA N= 4, AAA N=3, Control N= 3 to 5, Data are presented as the mean ± SD. P calculated with unpaired t-test. *P<0.05.

## Data Availability

All data produced in the present study are available upon reasonable request to the authors

## Acknowledgements

We want to thank Dr. Lea Davis for her technical assistance with the BioVU data set analysis. The Synthetic Derivative and BioVU projects at Vanderbilt University Medical Center are supported by numerous sources: institutional funding, private agencies, and federal grants, including the NIH funded Shared Instrumentation Grant S10OD017985 and S10RR025141; CTSA grants UL1TR002243, UL1TR000445, and UL1RR024975 from the National Center for Advancing Translational Sciences. Its contents are solely the responsibility of the authors and do not necessarily represent official views of the National Center for Advancing Translational Sciences or the National Institutes of Health. Genomic data are also supported by investigator-led projects that include U01HG004798, R01NS032830, RC2GM092618, P50GM115305, U01HG006378, U19HL065962, R01HD074711; and additional funding sources listed at https://victr.vumc.org/biovu-funding/. UNCF/BMS EE United Negro College Fund/Bristol-Myers Squibb E.E. Just Postgraduate Fellowship in the Life Sciences Fellowship to H.K.B., The United Negro College Fund/Bristol-Myers Squibb E.E. Just Faculty Fund, Burroughs Wellcome Fund Career Awards at the Scientific Interface Award, Burroughs Wellcome Fund Ad-hoc Award, National Institutes of Health Small Research Pilot Subaward to 5R25HL106365-12 from the National Institutes of Health PRIDE Program, DK020593, Vanderbilt Diabetes and Research Training Center for DRTC Alzheimer’s Disease Pilot & Feasibility Program to A.H.J. Research was supported by NHLBI Research Scientist Development Grant (1K01HL135461-01) to J.A.G. Small Research Project (SRP) Subaward from PRIDE-CVD to J.A.G, SRPs are funded by an NHLBI grant #R25 HL105446 to Dr. Mohamed Boutjdir.

## Author contributions

D.C., I.A., A.H.J., and J.A.G. conceived and designed research. D.D.H. and A.G.M. performed RNAseq data analysis. T.W.M., K.V.A, P.M.A., Q.S.W., H.K.B., D.F.D., N.J.C. provided access to BioVU data and performed the analysis. D.C., I.A., P.B.L and J.A.G. performed experiments, and Z.V., and A.G.M collected mouse samples. Human samples were collected by M.G.L., F.T.B., and J.A.C. The first draft of the manuscript was written by D.C., I.A., T.W.M., D.D.H. and J.A.G. All the authors listed revised, edited, and approved the manuscript.

## Institutional Review Board Statement

All studies involving human samples was IRB Board approved (Perioperative vascular reactivity; IRB Number: 191351).

## Supplemental Materials and Methods

### Animals

Six-week-old Myh11Cre^ERT2^/Sox6^fl/fl^ (Sox6-KO) and Myh11Cre^ERT2^/Sox6^wt/wt^ (Sox6-WT) control littermate mice were used in this study. The precise number of the animals for Sox6 KO and WT littermates are provided in the figure legends. The sequences of the primers for genotyping are: Sox6fl/fl, Forward Primer: 5’-GTC ACT CAG AGG TTA CTA TGG TG-3’; Reverse Primer: 5’-TTG GAG GCT TTA GCA GCT CTC-3’ ^1^. Myh11 Cre^ERT2 2–5^., Transgene Forward 5’-TGA CCC CAT CTC TTC ACT CC-3’, Transgene Reverse 5’-AGT CCC TCA CAT CCT CAG GTT-3’, Internal positive controls Forward 5’-CAG CCA ACT TTA CGC CTA GC-3’, and Internal positive control Reverse 5’-TCT CAA GAT GGA CCT AAT ACG G-3’. Band to each of the PCR products obtained are presented in Supplementary Figure 13A. The expression of Cre in aortic smooth muscle cells was determined using IHC with a protocol previously reported ^1, 6^ and presented in Supplementary Figure 13B. IHC was performed by following the previously published protocol of our lab^1, 6^. Briefly, aortas were fixed with 10% neutral buffered formalin solution, dehydrated in graduated ethanol series, and embedded in paraffin. Aorta sections were cut at 10 microns thickness. Histo-Clear solution (catalog no. HS-202, National Diagnostics) was used to deparaffinized the sections and permeabilized with 0.2% Triton X-100 at room temperature (RT). Thereafter, sections were blocked with 5% BSA-PBS at RT and incubated with primary antibodies prepared in 1% BSA-PBS overnight at 4 °C. Cre (Novus cat # NB100-56133F) and alpha smooth muscle actin (abcam cat # ab21027). Next morning, sections were washed with PBS (3X5 min). After three washes, sections were incubated with fluorochrome-conjugated secondary antibodies for 1 h at RT. The secondary antibodies were prepared in 1% BSA-PBS (1/500) and were chosen based on the primary antibodies and Alexa fluor fluorophores (ThermoFisher). DAPI was used to counterstain the nuclei. The tissue sections images were acquired with Nikon Eclipse Ti, (Software NIS-Elements AR 4.40.00 64-bit).

### Calculating genetically-regulated gene expression (GREX) in Vanderbilt University Medical Center biobank, BioVU

Vanderbilt University Medical Center curates a biorepository of genotype data matched to de-identified electronic health records (EHR) for over 259,000 individuals. Details on the BioVU program, including oversight, patient engagement and ethical considerations have been previously discussed ^7^. This opt-in program collects leftover blood samples from patient visits at clinics across Tennessee. BioVU initiated sample collection in 2007 and is ongoing. Data from the 1000 Genome Project were used to define genetic ancestry with principal component analysis (PCA) ^8, 9^. Genetically regulated gene expression was calculated in BioVU individuals from models built using the genotype-tissue expression (GTEx) project data ^10^. GTEx version 8 includes genotype data matched to RNA sequencing for over 838 donors across 49 distinct tissues. The best performing GREX models based on the highest r2 values from PrediXcan, UTMOST, and JTI approaches were used to model gene expression for SOX6 and additional genes from BioVU genotype data ^11–13^.

### GREX Phenotype association study in BioVU

Aortic aneurysm cases and controls were identified in BioVU by at least two mentions of the aortic aneurysm phecode (442.1) mapped from ICD9/10 diagnosis billing codes (International Classification of Diseases, 9th and 10th editions), including: aortic aneurysm and dissection, dissecting aneurysm of aorta, dissecting aneurysm of aorta-unspecified site, dissection of aorta thoracic, dissecting aneurysm of abdominal aorta, dissecting aneurysm of thoracoabdominal aorta, thoracic aortic aneurysm-ruptured, thoracic aneurysm without rupture, aortic aneurysm of unspecified site-ruptured, thoracoabdominal aortic aneurysm-ruptured, thoracoabdominal aortic aneurysm-ruptured, thoracoabdominal aortic aneurysm-without mention of rupture, aortic aneurysm-not otherwise specified, aneurysm of aorta in diseases classified elsewhere. Controls included individuals without any mention of the aortic aneurysm phecode and excluded individuals with any mention of the similar, yet distinct phecodes: diseases of arteries, arterioles, and capillaries identified using the PheWAS package in R (version 0.99.5-2 and 3.6.0, respectively) ^14, 15^. MultiXcan was used to combine the GREX of each gene across all tissues ^16^.

### GREX Laboratory-wide association scan (LabWAS) in BioVU

Laboratory values were extracted from the Vanderbilt University Medical Center de-identified EHR database and cleaned using the QualityLab pipeline as previously described ^17^.

### Aortic aneurysm diagnosis LabWAS in the Vanderbilt Synthetic Derivative

The Vanderbilt Synthetic Derivative is a de-identified database of the electronic health records for over 3 million patients at Vanderbilt, dating back more than 30 years ^18^.

### RT-qPCR

Preparation of template cDNA. Aortic Aneurysm Aorta (AAA), Thoracic Aorta Aneurysm (TAA) and Control Aorta (CA) samples were collected from the OR at Vanderbilt University Medical Center, flash freeze in liquid nitrogen, and stored at −80°C. The RNA was extracted from a portion of each sample by homogenizing two times during 30 seconds with Tissue-Tearor (Model 985370, BioSpec Products, Inc.) resuspended in 600 uL RNA Lysis Buffer from Quick-RNA MiniPrep kit (ZYMO RESEARCH) RNA was extracted following the manufacturer’s instructions. The amount of RNA extracted was quantified on NanoDrop One (Thermo Scientific). The cDNA was obtained from the RNA extracted from the samples using SuperScript VILO MasterMix (Invitrogen, ThermoFisher Scientific) following manufacturer’s instructions. The amount, quality and purity of the cDNA was measured using the NanoDrop One (Thermo Scientific). Real-time PCR was performed in the QuantStudio 3 Real-Time PCR System (Applied Biosystems) using TaqMan Fast Advanced Master Mix (Applied Biosystems, containing AmpliTaq Fast DNA Polymerase, uracil-N-glycosylase, dNTPs with dUTP, ROX dye as passive reference, and optimized buffer components). The total reaction volume (20 μL) consisted of the following: 1 μL cDNA (100 ng), 10 μL TaqMan Fast Advanced Master Mix, 1 μL of each TaqMan Gene Expression Assay, and 8 μL ultrapure DNase-free water. The thermal cycle parameters were as follows: UNG incubation at 50°C for 2 min, polymerase activation at 95°C for 20 s, denaturation at 95°C for 1 s and then annealing and extension at 60°C for 20 s. We used Thermofisher Taqman proves for each gene assay. The gene assays used in this study were polycystin 1 (PKD1), gremlin 1 (GREM1), coactosin like F-actin binding protein 1 (COTL1), joining chain of multimeric IgA and IgM (JCHAIN), C-C motif chemokine ligand 21 (CCL21), ATPase H+ transporting accessory protein 2 (ATPGAP2), tumor necrosis factor alpha (TNFα), transforming growth factor beta (TGFβ1), SRY-box transcription factor 6 (SOX6), matrix metallopeptidase 9 (MMP9), matrix metalloproteinase (MMP2), and glyceraldehyde-3-phosphate dehydrogenase (GAPDH) and eukaryotic translation initiation factor 2B subunit alpha (EIF2B1) were used as housekeeping genes. Real-Time qPCR data normalization and analysis. The samples included in the study allowed to see how normally the gene assays are expressed under tissue normal conditions, in the other hand the pathological conditions allowed us to see if it has any difference in terms of expression over those candidate genes. The comparative C_T_ method was used to calculate the relative expression of the transcripts in all the samples and genes were normalized according to the housekeeping genes.

### Western blot

Preparation of BCA Standard Curve and getting protein concentration from the samples. The BCA standard curve was made on a microplate and constructed using the Pierce™ BCA Protein Assay Kit (Thermo Scientific) following manufacturer’s instructions mixing the reagent A and reagent B (50:1) with the BCA and dilutions, and with the samples (AAA, TAA, and CA) from protein extraction procedure. Incubation was performed during 30 minutes at 37 °C, let the plate cool down and measure the absorbance at 562nm on a plate reader. With the results, the standard curve was constructed and calculated the original concentration of the samples to define how much amount of protein is going to be added to the electrophoresis gel. SDS-PAGE Electrophoresis Gel. To perform the electrophoresis the Mini-PROTEAN TGX Precast Gels (BIO-RAD, Cat. #456-1096) were used once the running buffer 25 mM Tris, 190 mM Glycine, and 0.1% SDS) was added to the electrophoresis chamber Mini-PROTEAN Tetra System (BIO-RAD). The samples were prepared with 10 μL 4x Laemmli Sample Buffer (BIO-RAD, Cat. #1610747) and 10 μL of sample and Nuclease free water. The samples were heated during 5 min at 95 °C and each well was loaded with 30 ug of protein with 10 μL of Precision Plus Protein Standards (BIO-RAD) ladder. The conditions to run the gels were stablished for 5 min at 50V and then for 70 min at 100 V. The sponges and filter papers were incubated in cold transfer buffer (25 mM Tris, 190 mM Glycine, and 20% methanol) for 15 min at 4 °C. The PVDF membrane was activated in methanol for 30 seconds, put between the electrodes and the filter papers along with the polyacrylamide gel to run it for 120 min at 100 V in a 4 °C room. Once the electro transfer is completed, we can proceed with the antibodies looking for the protein of interest. Immunoblotting. The PVDF membrane is transferred directly in fresh blocking buffer (milk 5% in TBS 1X-T 0.1%) for 1-2 hours at room temperature, later the primary antibody is diluted (1:100) in milk 5% with TBS 1X-T 0.1% and it’s added to the PVDF membrane overnight at 4 °C, then washed 3x with TBS 1X-T 0.1% incubated for 10 min at room temperature. The second antibody is added (1:1000) diluted in milk 5% with TBS 1X-T 0.1%, let incubate it for 1 hour at room temperature and washed 3x with TBS 1X-T 0.1% incubated for 10 min at room temperature. Now revealed with Clarity Western ECL Substrate (BIO-RAD) following software’s instructions. For stripping the blot Restore Western Blot Stripping Buffer (Thermo Scientific) was used and the immunoblotting was repeated if necessary, according to the different antibodies needed. Protein bands were quantified and normalized with the house-keeping gene beta actin (Sigma-Aldrich cat # A1978) using software integrated with the image station.

### RNA Sequencing

RNA was isolated from Aortic Aneurysm Aorta (AAA), Thoracic Aorta Aneurysm (TAA) and Control Aorta (CA). Total RNA was isolated using a PicoPure Arcturus kit (Invitrogen) according to the manufacturer’s instructions. Complimentary cDNA was generated with an Ovation Pico WTA System V2 kit (NuGEN) which maintains the stoichiometry of the original RNA population. library construction was performed NEBNext® Ultra™ II for DNA Library Prep. All samples are sequenced and performed at multiplex Paired-End 150 bp on the Illumina NovaSeq 6000 by VANTAGE core at VUMC. Raw sequencing data was uploaded and analyzed using the Illumina BaseSpace platform. Sequences (151 bp paired end) were trimmed from both ends to 125 bp using the FastQ Toolkit app. Transcripts were aligned to the human hg19 genome and quantified with the RNA Express app. Differentially expressed genes (DEGs) were determined using a false discovery rate (FDR) threshold of 0.05 and expression mean count >10 for any group. Data have been deposited into the GEO database (accession GSE202267). Expression heatmaps were generated with Morpheus (Broad Institute) via hierarchical clustering using average linkage and Spearman’s correlation. Enriched pathway predictions were performed with Ingenuity Pathway Analysis (IPA, QIAGEN) and through gene set enrichment analysis (GSEA, WebGestalt.org). An absolute activation Z-score of >;2.0 and a p-value of <0.05 were deemed as significant for enriched terms.

### Human and murine tissue staining

Abdominal aortas of anesthetized (isoflurane) mice 4 weeks after Ang II fusion were harvested and fixed overnight at 4 °C in 4%v/v neutral buffered formalin, followed by incubation in 30%w/v sucrose for a further 24hrs at 4 °C. The aortas were paraffin embedded. For each section, 5 10-micron slices were taken and mounted onto a glass slide. Masson’s trichrome, Hematoxylin and Eosin, and Van Geison staining was then performed according to standard techniques in the Pathology core at VUMC. Human samples were fixed overnight at 4 °C in 4%v/v neutral buffered formalin, followed by incubation in 30%w/v sucrose for a further 24hrs at 4 °C. The aortas were paraffin embedded. For each section, 5 10-micron slices were taken and mounted onto a glass slide. Masson’s trichrome, Hematoxylin and Eosin, and Van Geison staining was then performed according to standard techniques in the Pathology core at VUMC.

## Supplemental Figures

**Supplemental Figure 1.**
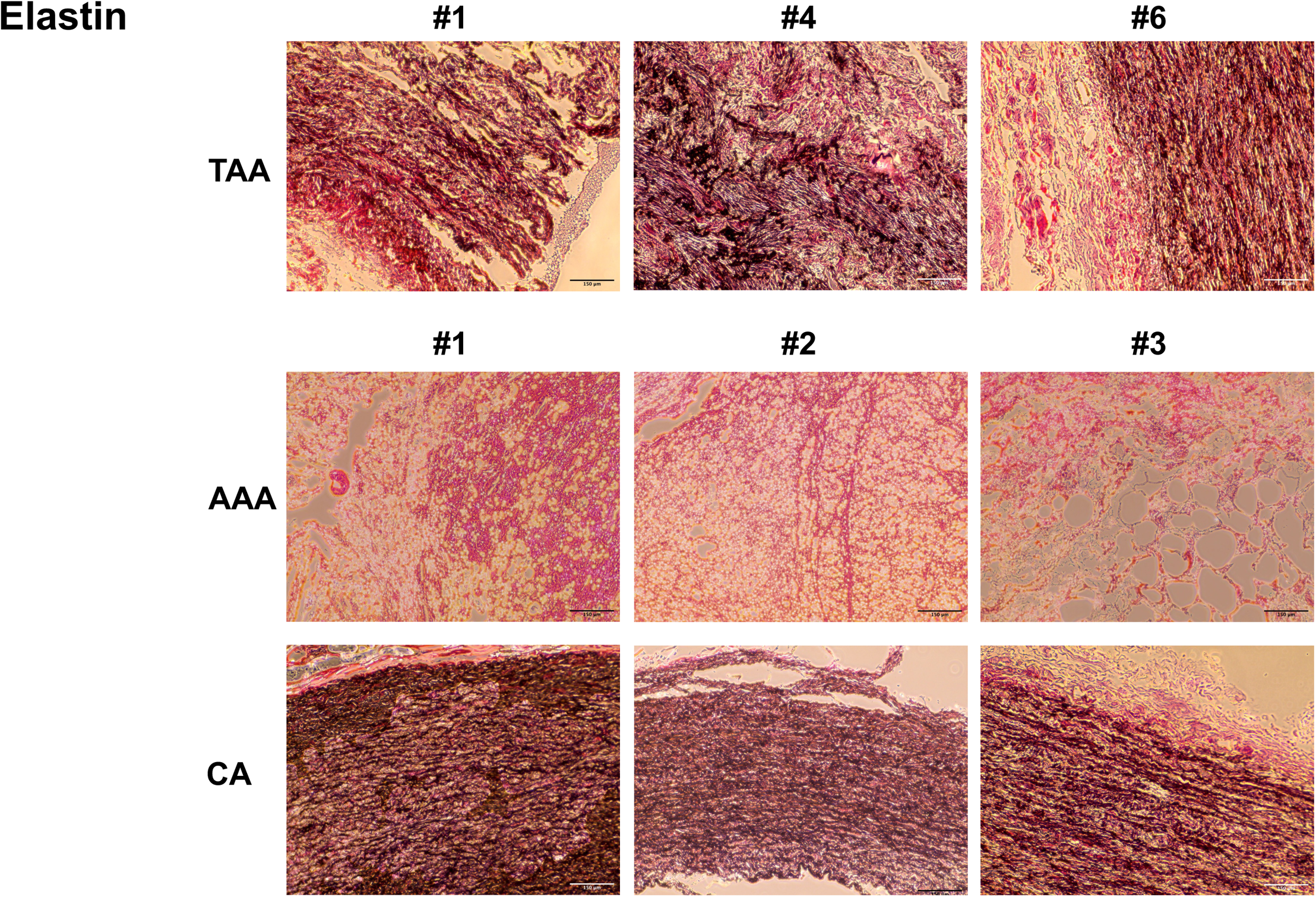
Human abdominal and Thoracic aortic aneurysm elastin staining. Aortic Aneurysm Aorta (AAA), Thoracic Aorta Aneurysm (TAA) and Control Aorta (CA) samples were collected from the OR at Vanderbilt University Medical Center. Full informed consent was obtained for all tissue samples. Tissue was fixed in 4% formalin at 4 degrees Celsius overnight. Tissue was processed and stained in the Pathology Core VUMC.

**Supplemental Figure 2.**
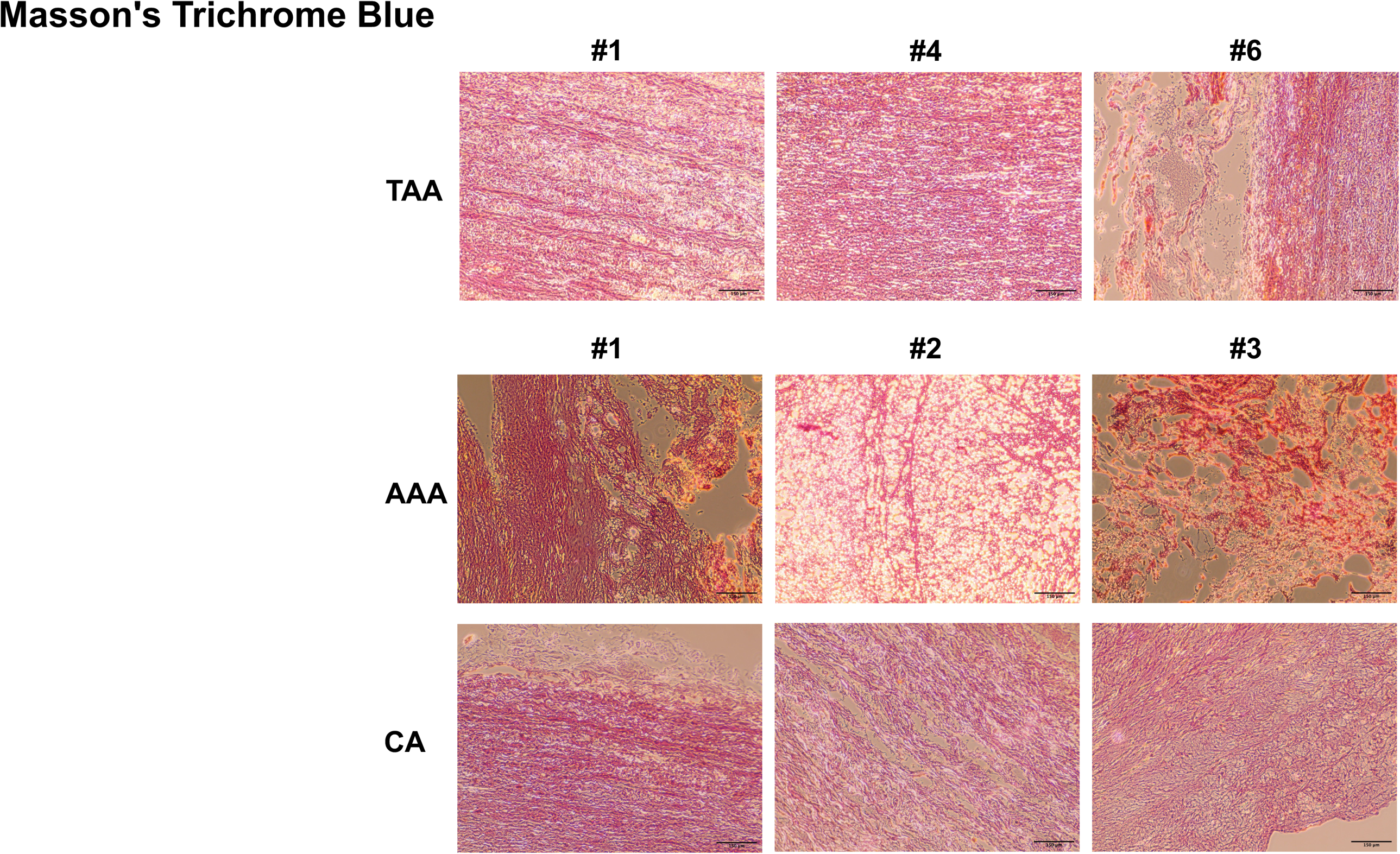
Human abdominal and Thoracic aortic aneurysm elastin staining. Aortic Aneurysm Aorta (AAA), Thoracic Aorta Aneurysm (TAA) and Control Aorta (CA) samples were collected from the OR at Vanderbilt University Medical Center. Full informed consent was obtained for all tissue samples. Tissue was fixed in 4% formalin at 4 degrees Celsius overnight. Tissue was processed and stained in the Pathology Core VUMC.

**Supplemental Figure 3.**
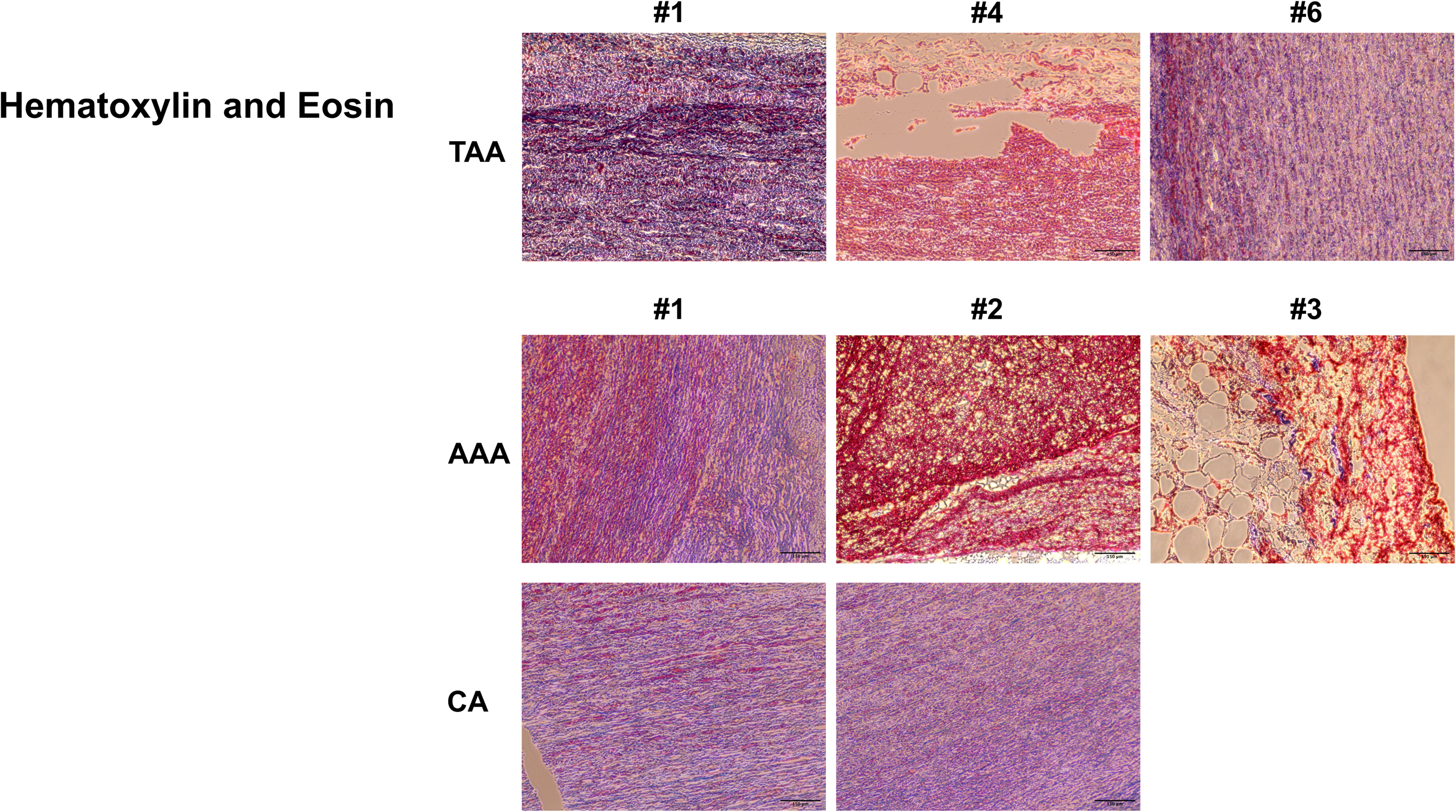
Human abdominal and Thoracic aortic aneurysm elastin staining. Aortic Aneurysm Aorta (AAA), Thoracic Aorta Aneurysm (TAA) and Control Aorta (CA) samples were collected from the OR at Vanderbilt University Medical Center. Full informed consent was obtained for all tissue samples. Tissue was fixed in 4% formalin at 4 degrees Celsius overnight. Tissue was processed and stained in the Pathology Core VUMC.

**Supplemental Figure 4.**
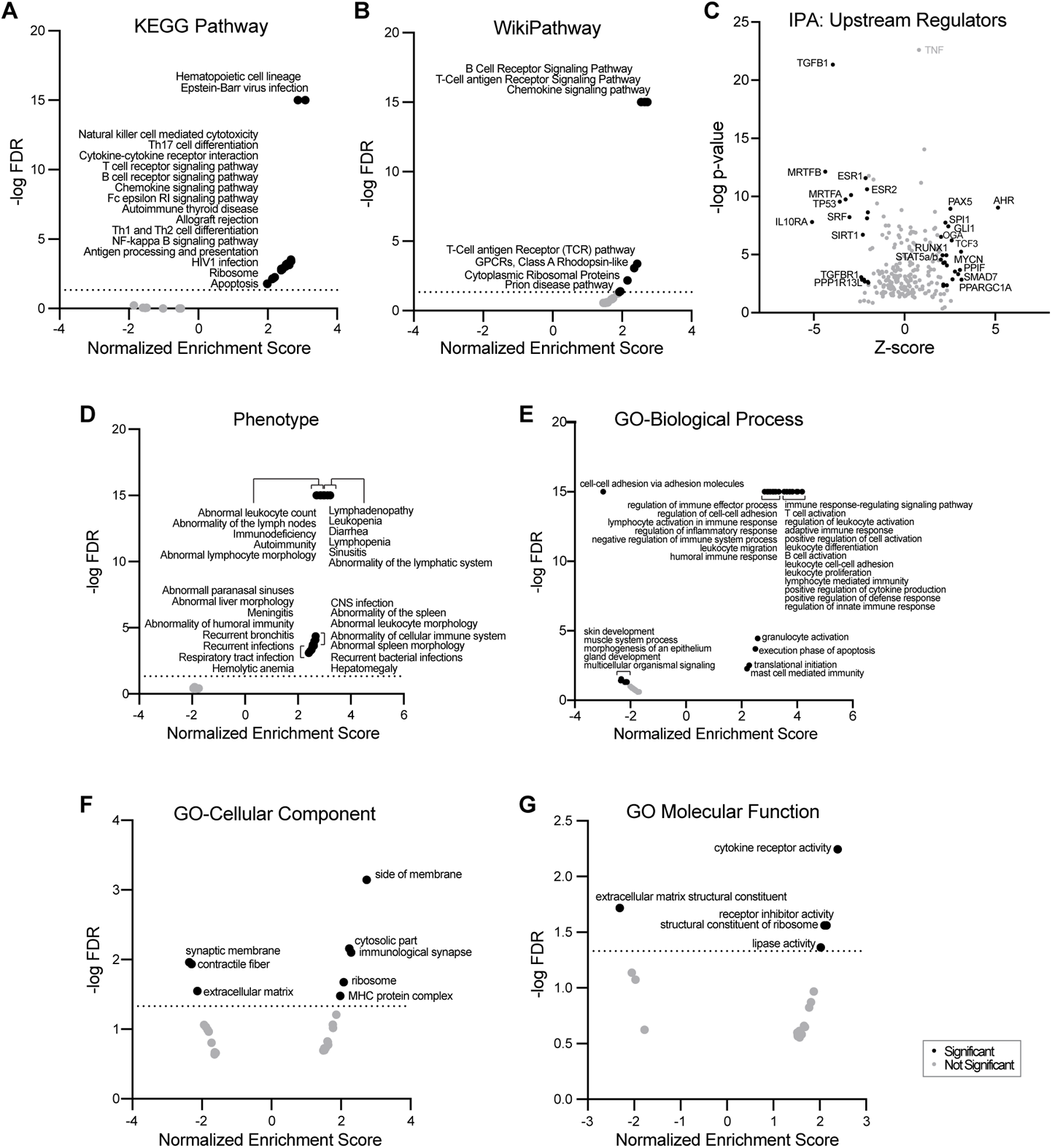
Human abdominal aortic aneurysm RNA sequence tissue analysis. Enrichment analyses of significant DEGs found in abdominal aortic aneurysm (AAA) samples by RNA-sequencing. DEGs were analyzed by gene set enrichment analysis (GSEA, panels A, B, D-G) or by Ingenuity pathway analysis (IPA, panel C). Type of enrichment analysis are indicated for each panel. GO, gene ontology. Enriched terms meeting a threshold p-value <0.01 (C) or false discovery rate (FDR) <0.05 (dotted lines in A, B, D-G), and an absolute normalized enrichment score or Z-score of >2.0 were deemed as significant (black circles). Non-significant points are marked as gray circles.

**Supplemental Figure 5.**
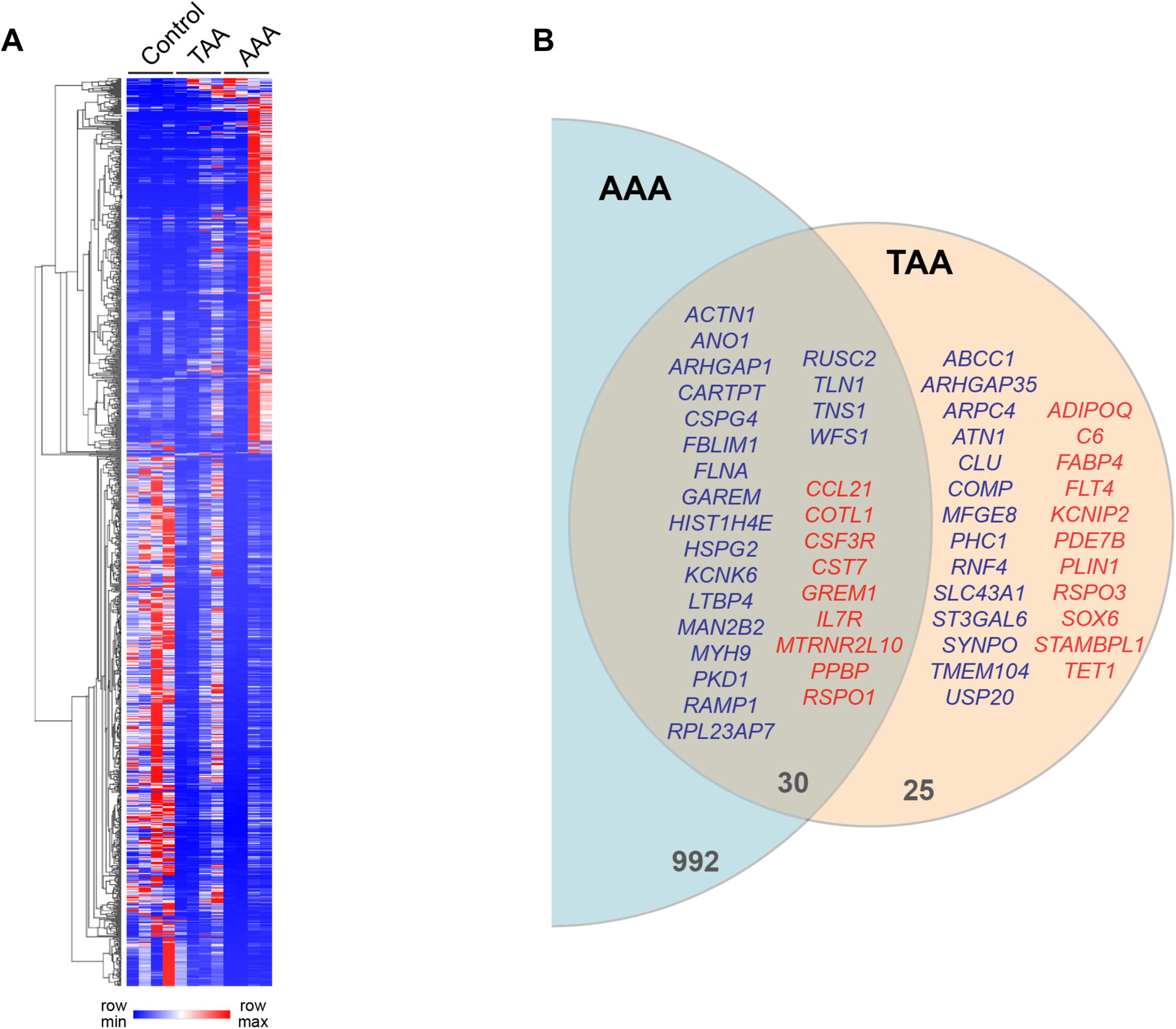
Human abdominal aortic aneurysm and Thoracic aortic aneurysm RNA sequence tissue analysis. **(A)** Heatmap of 1022 and 55 significant differentially expressed genes (DEGs) found in AAA and TAA samples relative to control respectively with 30 genes in common (n=4 per group). **(B)** Venn diagram showing the common genes between AAA and TAA with upregulated genes in read and down regulated genes in blue.

**Supplemental Figure 6.**
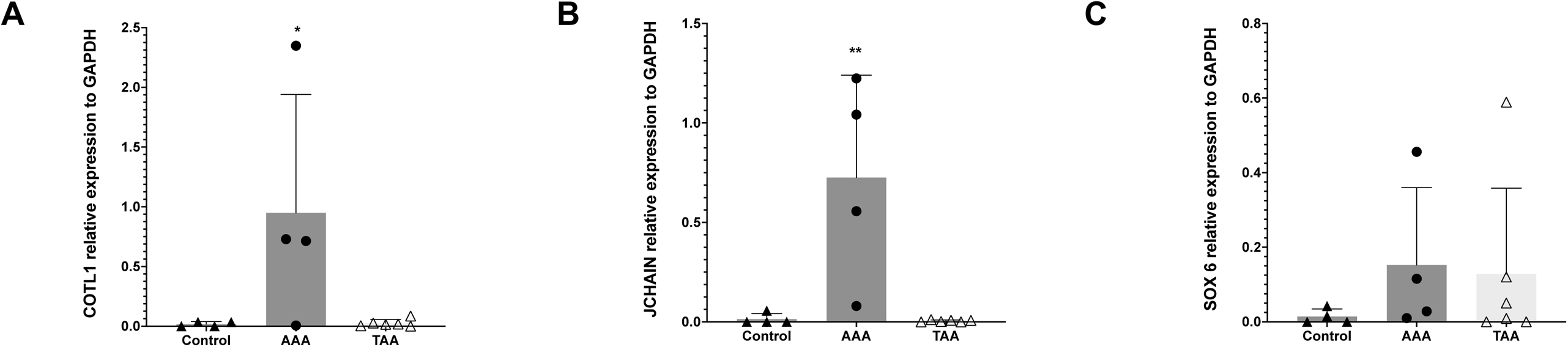
Human abdominal and Thoracic aortic aneurysm mRNA quantification. Abdominal Aortic Aneurysm (AAA), Thoracic Aorta Aneurysm (TAA) and Control Aorta (CA) samples were collected from the OR at Vanderbilt University Medical Center. Full informed consent was obtained for all tissue samples. RNAseq validation of upregulated genes. **(A)** COTL1. **(B)** JCHAIN. **(C)** SOX6. Relative expression to GAPDH. TAA N=6, AAA N=4, Control N= 4, Data are presented as the mean + SD. P calculated with unpaired t-test. *P<0.05. **P<0.01.

**Supplemental Figure 7.**
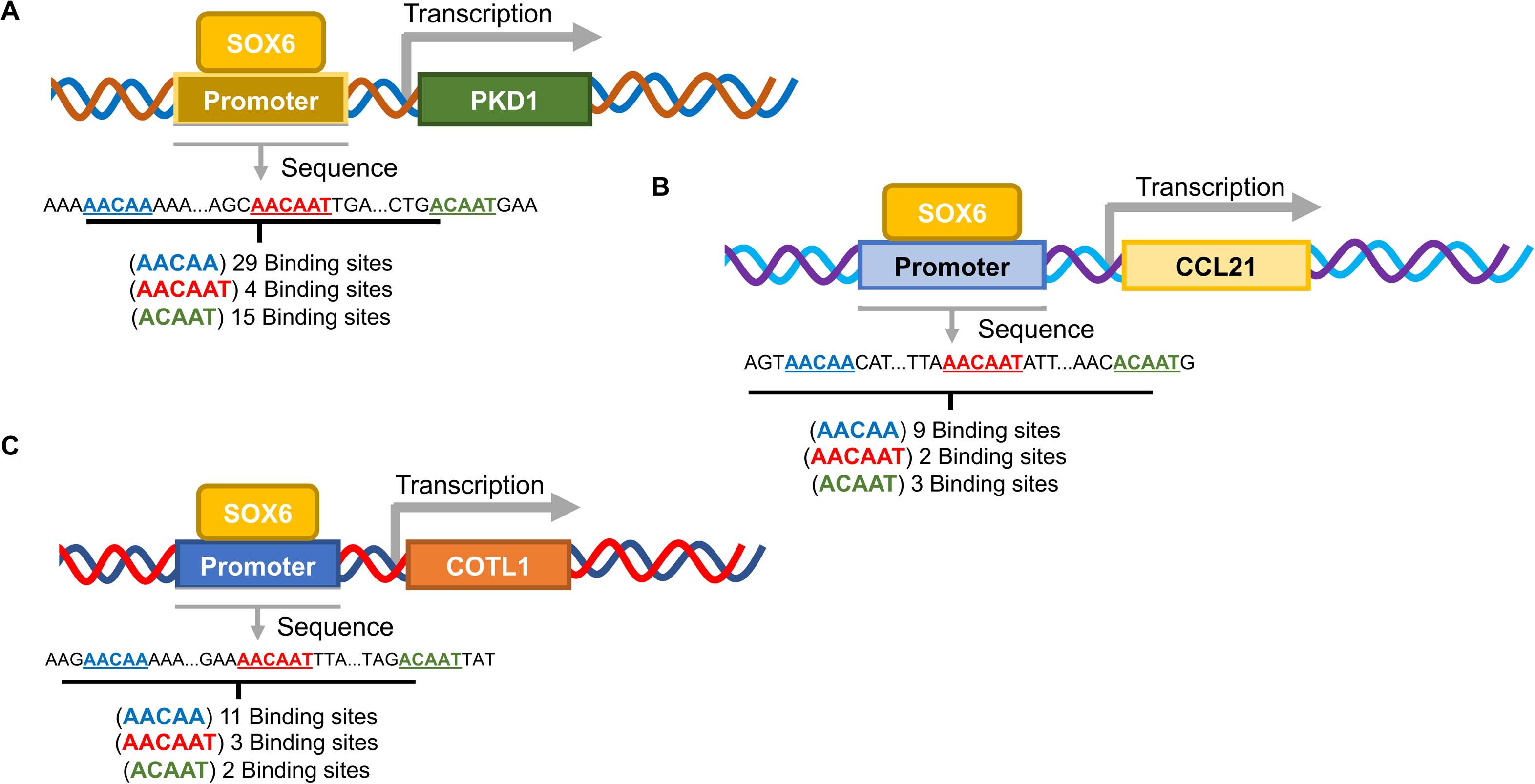
SOX6 binding sites in the promoters of PKD1, CCL21 and COTL1. The *in-silico* analysis of upstream region from the start codon of the genes of interest, looking for the SOX6 DNA binding canonical sequences (6-5 bp). The *in-silico* analysis shows several potential binding sites for SOX6. **(A)** shows the number of sites predicted for the PKD1 gene, **(B)** for the CCL21 gene, and **(C)** for the COTL1 gene.

**Supplemental Figure 8.**
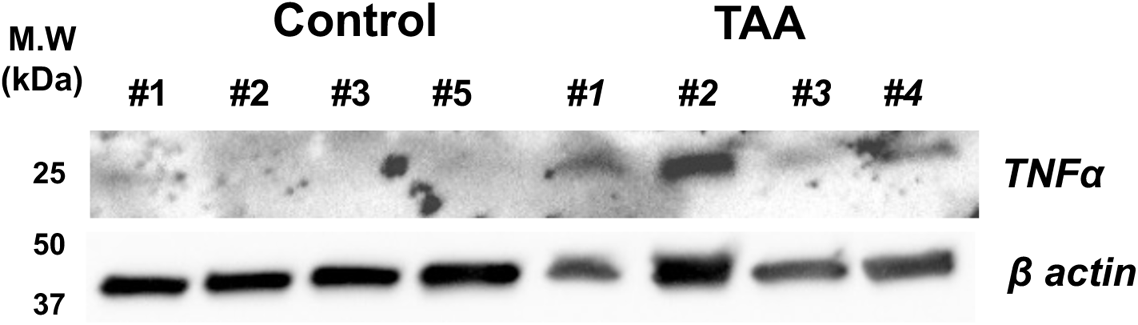
Human Thoracic aortic aneurysm TNFα expression Western blot analysis. Human aorta samples were collected from tissue discarded after TAA or coronary artery bypass surgery (control). Samples were flash freeze in liquid nitrogen and stored at −80°C. Expression of TNFα was analyzed by Western blot using a specific antibody. Representative Western blots images showing levels of TNFα expression in human TAA. Beta-actin was used as a loading control.

**Supplemental Figure 9.**
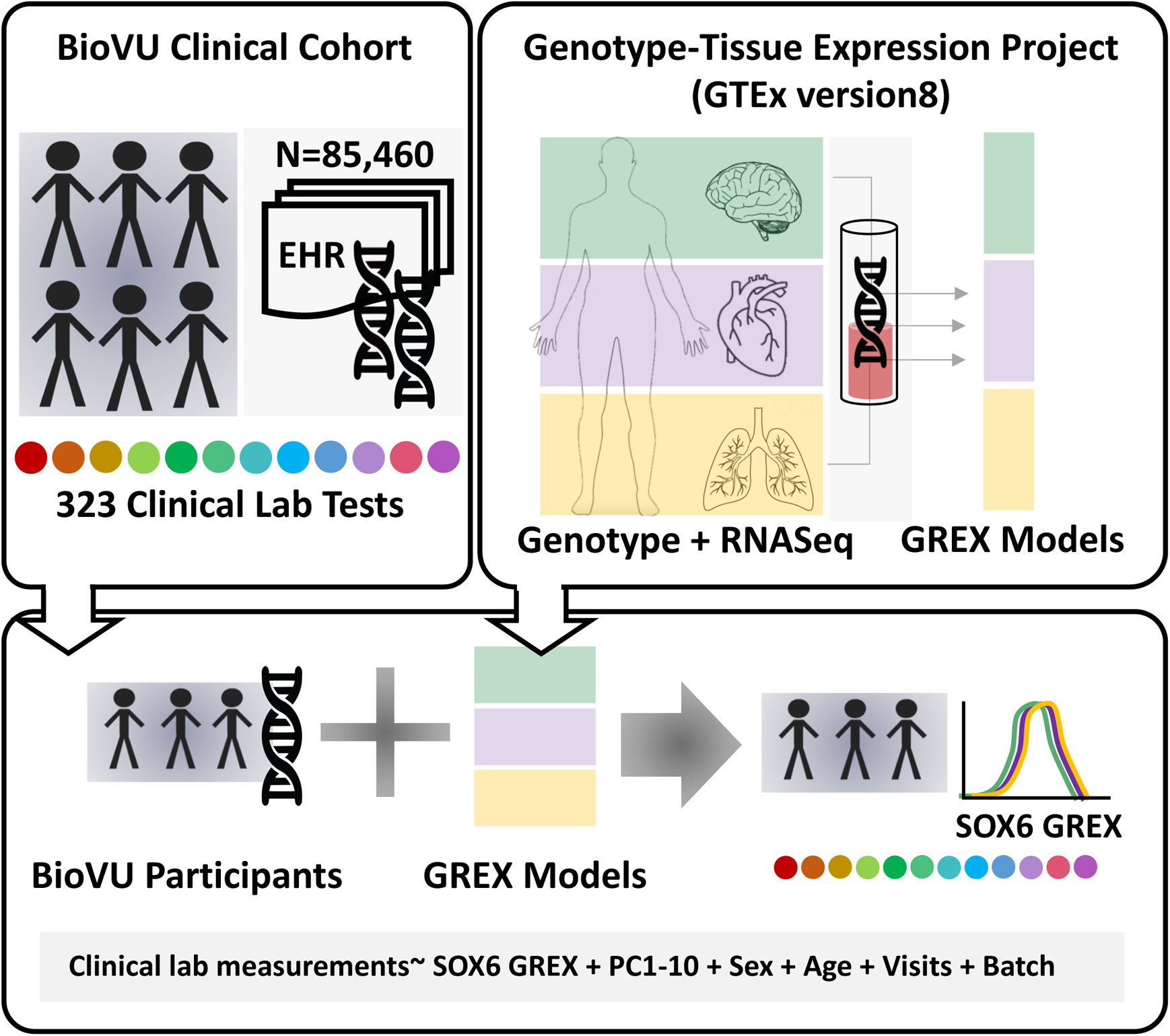
Clinical lab-wide scan for SOX6 genetically-regulated gene expression (GREX) in a medical biobank. Clinical lab values from BioVU participants were extracted from Vanderbilt’s de-identified electronic health record database (n= 85,460, top left panel). Genetically-regulated gene expression for SOX6 and SOX6-regulated genes were calculated in BioVU participants using models built from the GTEx version 8 data (top right panel), which contains genotype data matched to RNA-Seq data from 838 donors across 49 tissues. Imputed gene expression was calculated and tested for association across up to 323 clinical lab tests using linear regression models (bottom panel), accounting for genetic ancestry (principle components/PC 1-10, sex, age, number of medical center visits, and genotyping batch).

**Supplemental Figure 10.**
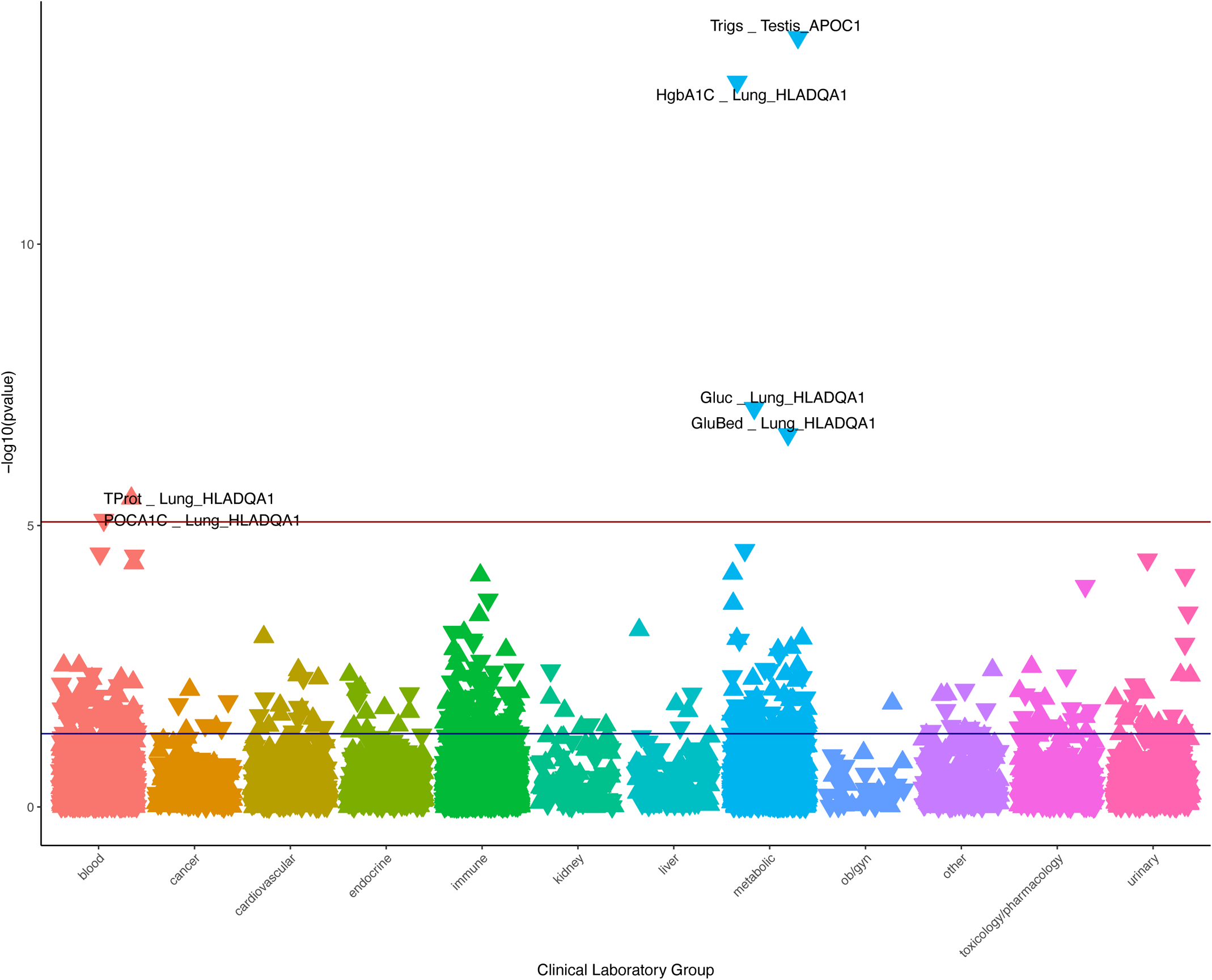
Clinical lab-wide scan for AAA/TAA genetically-regulated gene expression (GREX) in individuals of European ancestry. Clinical lab values from BioVU participants were extracted from Vanderbilt’s de-identified electronic health record database and genetically-regulated gene expression for SOX6 was calculated in BioVU participants of European ancestry (n=70,337). The GREX for SOX6 and SOX6-regulated genes was tested for association across 323 clinical lab tests using linear regression models, accounting for genetic ancestry (principle components/PC 1-10, sex, age, number of medical center visits, and genotyping batch). Associations that met the Bonferroni-corrected threshold (red line, p < 8.627×10-6) are labeled with lab name, GREX gene, and GREX tissue. Blue line represents p = 0.05.

**Supplemental Figure 11.**
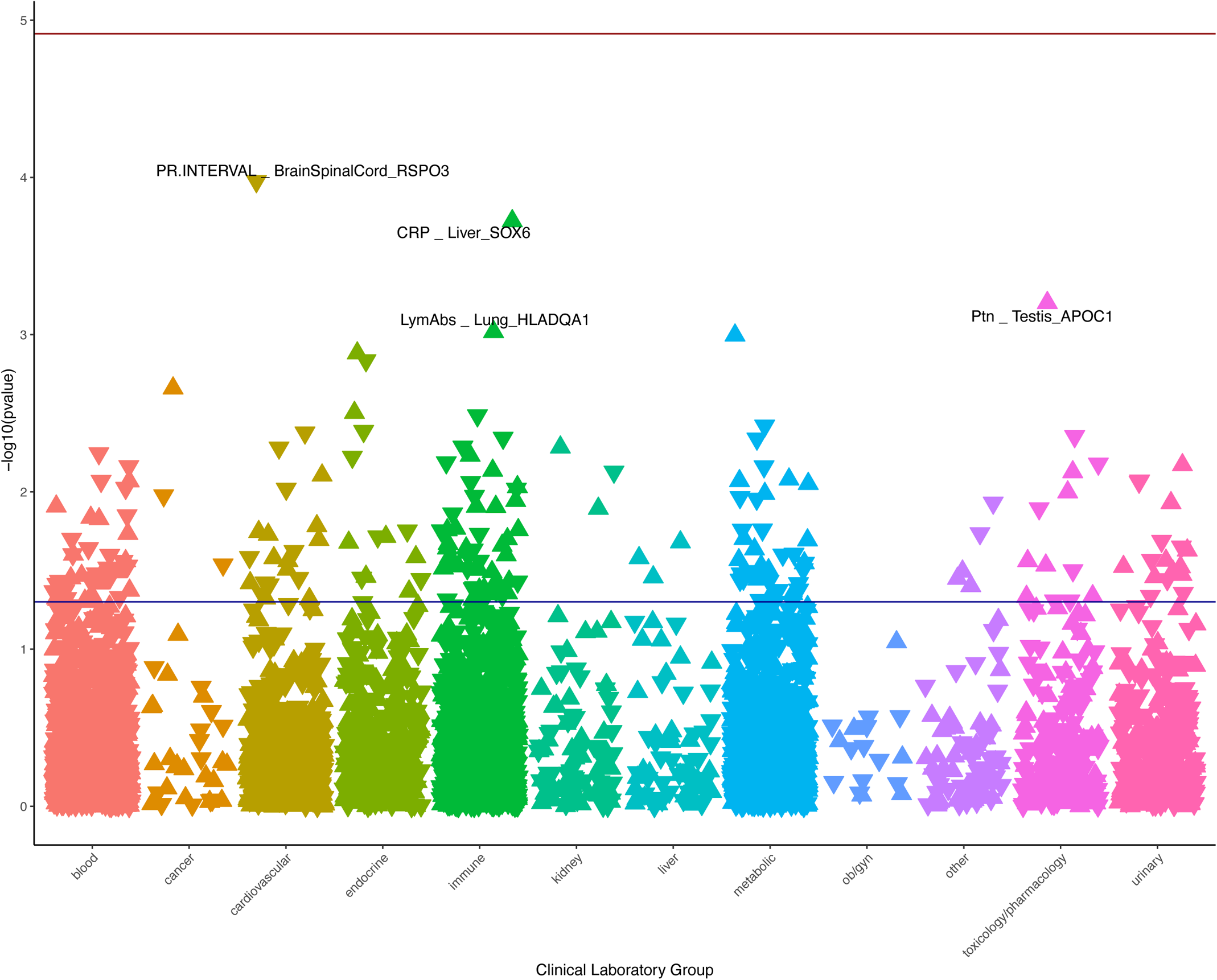
Clinical lab-wide scan for AAA/TAA genetically-regulated gene expression (GREX) in individuals of African ancestry. Clinical lab values from BioVU participants were extracted from Vanderbilt’s de-identified electronic health record database and genetically-regulated gene expression for SOX6 was calculated in BioVU participants of African ancestry (n=15,123). The GREX for SOX6 and SOX6-regulated genes was tested for association across 241 clinical lab tests using linear regression models, accounting for genetic ancestry (principle components/PC 1-10, sex, age, number of medical center visits, and genotyping batch). Associations that met the Bonferroni-corrected threshold (red line, p < 1.220×10^−5^) are labeled with lab name, GREX gene, and GREX tissue. Blue line represents p = 0.05.

**Supplemental Figure 12.**
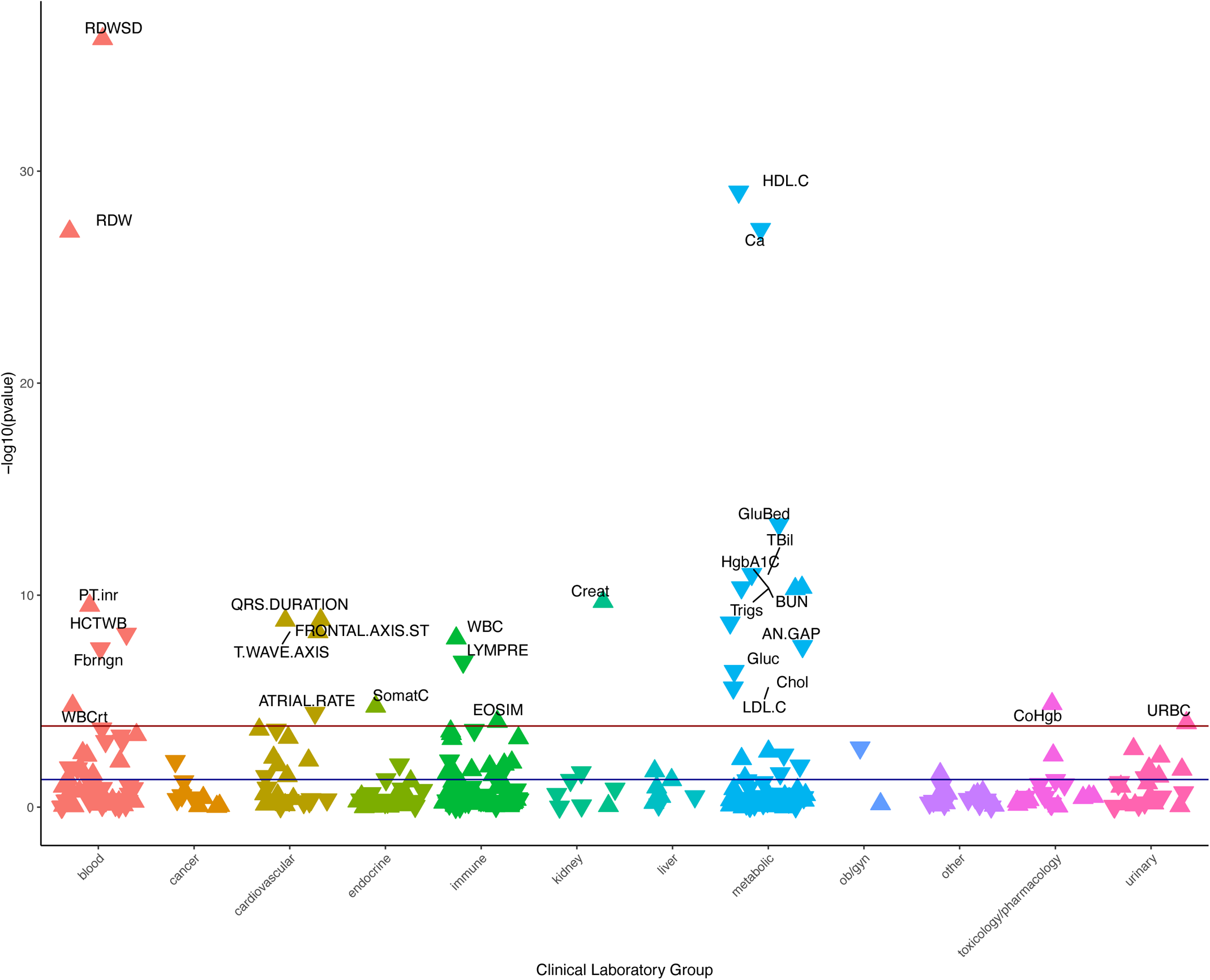
Clinical lab-wide scan for aortic aneurysm diagnosis. Clinical lab values were extracted from Vanderbilt’s de-identified electronic health record database for cases and controls of aortic aneurysm (n=2,273 and 1,268,204 respectively). The aortic aneurysm diagnosis was tested for association across 335 clinical lab tests using linear regression models, accounting for EHR-reported race and ethnicity, sex, age, and number of medical center visits). Associations that met the Bonferroni-corrected threshold (red line, p < 1.493 x 10^−4^) are labeled with lab name. Blue line represents p = 0.05.

**Supplemental Figure 13.**
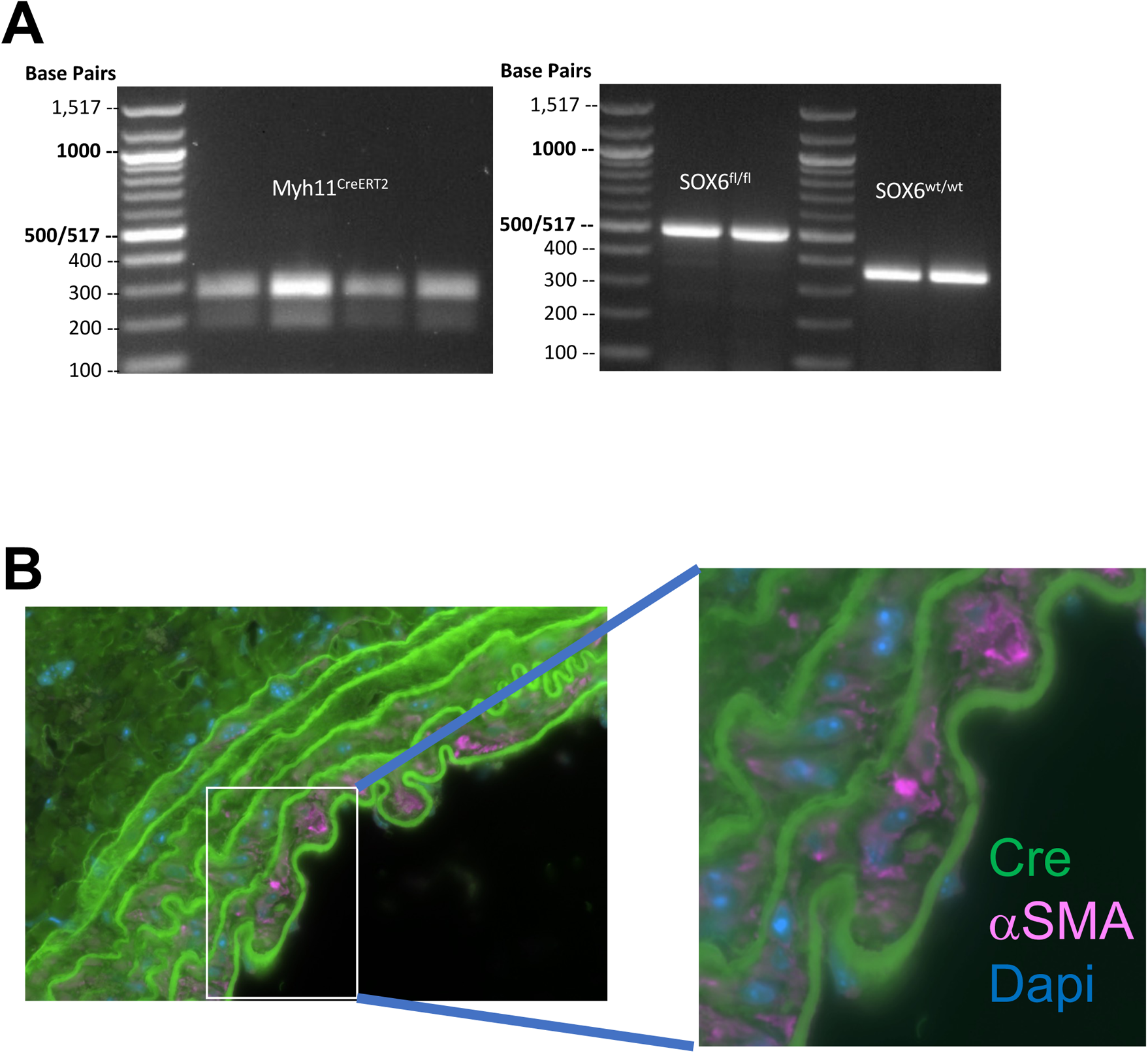
Myh11Cre^ERT2^/Sox6^fl/fl^ mice. **(A)** Mice Myh11Cre^ERT2^ and Sox6 PCR genotype products. **(B)** Myh11Cre^ERT2^/Sox6^wt/wt^ (Sox6 WT) mice were intraperitoneally with injected 2.5 mg/mouse of tamoxifen for 5 consecutive days. After Cre induction by tamoxifen abdominal aorta was harvested and stained to determine the Cre expression in smooth muscle cells. Cre (green) and αSMA (magenta). Smooth muscle cells labeled with alpha smooth muscle actin (αSMA). Autofluorescence in green from elastin fibers.

