## Supplemental Table 1 for "Sox6 expression and aneurysms of the thoracic and abdominal aorta"

**Supplemental Table 1. Control, TAA and AAA samples age and gender.**

| Sample ID | Age | Gender |
|---|---|---|
| Control # 1 | 51-55 | Male |
| Control # 2 | 66-70 | Female |
| Control # 3 | 61-65 | Female |
| Control # 4 | 61-65 | Male |
| TAA #1 | 46-50 | Male |
| TAA #2 | 81-85 | Male |
| TAA #3 | 51-55 | Male |
| TAA #4 | 71-75 | Male |
| AAA #1 | 61-65 | Male |
| AAA #2 | 61-65 | Male |
| AAA #3 | 56-60 | Male |
| AAA #4 | 66-70 | Male |

Control nonaneurysmal aortic tissue samples from ascending aorta punch sites for saphenous vein grafts from patients undergoing coronary artery bypass surgery. Thoracic aortic aneurysm (TAA). Abdominal aortic aneurysm (AAA). Age Range in years.
