## Supplemental Tables 3 and 4 for "Sox6 expression and aneurysms of the thoracic and abdominal aorta"

**Supplemental Table 3. Relative expression to GAPDH.**

| Relative expression to GAPDH | | | |
|---|---|---|---|
| **Gene** | **Control** | **AAA** | **TAA** |
| **SOX6** | 0.0144282 ± 0.0086424 | 0.1523593 ± 0.0898531 | 0.1281368 ± 0.0858647 |
| **PKD1** | 0.0294421 ± 0.0237806 | 0.1582278 ± 0.0703012 | 0.0647234 ± 0.0262019 |
| **GREM1** | 0.4811858 ± 0.2406540 | 0.1585224 ± 0.0959548 | 0.1829259 ± 0.1173131 |
| **COTL1** | 0.0177891 ± 0.0088954 | 0.9490997 ± 0.429449 | 0.0247907 ± 0.0115873 |
| **JCHAIN** | 0.014151 ± 0.0122551 | 0.7255081 ± 0.2227829 | 0.0047918 ± 0.001811 |
| **CCL21** | 0.0062402 ± 0.0054042 | 0.8569801 ± 0.4888084 | 0.0290346 ± 0.0114249 |
| **ATPGAP2** | 0.0475185 ± 0.0411522 | 0.9462023 ± 0.6826119 | 0.0472063 ± 0.0269386 |
| **TNF** | 0.0027019 ± 0.0023399 | 0.0101974 ± 0.006079 | 0.0019703 ± 0.0010461 |
| **TGFB1** | 0.1343831 ± 0.0843787 | 0.7155584 ± 0.2798296 | 0.1290861 ± 0.0820674 |
| **MMP9** | 0.0289058 ± 0.0213288 | 0.3620202 ± 0.1810107 | 0.0311398 ± 0.0119489 |
| **MMP2** | 0.3453867 ± 0.2258334 | 1.1536191 ± 0.4997104 | 0.145761 ± 0.0563883 |

Human Control Aorta, Abdominal Aortic Aneurysm (AAA), and Thoracic Aorta Aneurysm (TAA) samples were collected from the OR at Vanderbilt University Medical Center. Full informed consent was obtained for all tissue samples. RT-qPCR validation of RNAseq upregulated genes. Relative expression to GAPDH. TAA N=6, AAA N=4, Control N= 4, Data are presented as the mean + SD. P calculated with ANOVA. *P<0.05.

**Supplemental Table 4. Relative expression to EIF1B2.**

| Relative expression to EIF1B2 | | | |
|---|---|---|---|
| **Gene** | **Control** | **AAA** | **TAA** |
| **SOX6** | 0.6691274 ± 0.579481 | 18.7454412 ± 8.5936152 | 1.8582904 ± 0.7778931 |
| **PKD1** | 4.9693764 ± 3.9689996 | 4.1942074 ± 1.9362906 | 0.6684659 ± 0.2261999 |
| **GREM1** | 0.7619918 ± 0.5388096 | 2.5122468 ± 1.950472 | 2.0191594 ± 0.9141089 |
| **COTL1** | 0.8991971 ± 0.6732272 | 0.6235681 ± 0.23706 | 0.4426157 ± 0.1680298 |
| **JCHAIN** | 0.500238 ± 0.4332188 | 0.7276087 ± 0.2579229 | 0.1523504 ± 0.0578873 |
| **CCL21** | 1.1343858 ± 0.9824069 | 1.0043561 ± 0.4038473 | 0.7475581 ± 0.3049075 |
| **ATPGAP2** | 0.1489707 ± 0.1290124 | 9.122103 ± 6.4991056 | 0.9295919 ± 0.3193484 |
| **TNF** | 2.6199542 ± 2.2689469 | 21.3076284 ± 11.0560302 | 0.04651219 ± 0.0170214 |
| **TGFB1** | 1.3228938 ± 0.9021874 | 0.796285 ± 0.3173656 | 1.1964455 ± 0.4894253 |
| **MMP9** | 2.3766728 ± 1.7594192 | 0.6549987 ± 0.4302063 | 0.4464075 ± 0.1813713 |
| **MMP2** | 3.0240608 ± 2.5031633 | 0.5615069 ± 0.1977585 | 2.5800763 ± 1.2777419 |

Human Control Aorta, Abdominal Aortic Aneurysm (AAA), and Thoracic Aorta Aneurysm (TAA) samples were collected from the OR at Vanderbilt University Medical Center. Full informed consent was obtained for all tissue samples. RT-qPCR validation of RNAseq upregulated genes. Relative expression to EIF1B2. TAA N=6, AAA N=4, Control N= 4, Data are presented as the mean + SEM. P calculated with ANOVA. *P<0.05
